# COVCOG 1: Factors predicting Cognitive Symptoms in Long COVID. A First Publication from the COVID and Cognition Study

**DOI:** 10.1101/2021.10.26.21265525

**Authors:** Panyuan Guo, Alvaro Benito Ballesteros, Sabine P Yeung, Ruby Liu, Arka Saha, Lyn Curtis, Muzaffer Kaser, Mark P Haggard, Lucy G Cheke

**Affiliations:** Department of Psychology, University of Cambridge; Department of Psychiatry, University of Cambridge; School of Psychology, College of Life and Environmental Sciences, University of Exeter

**Keywords:** Long COVID, Cognition, Neurological, Memory, Executive Functions, Language, COVID-19, Symptoms

## Abstract

Since its first emergence in December 2019, coronavirus disease 2019 (COVID-19), caused by severe acute respiratory syndrome coronavirus 2 (SARS-CoV-2), has evolved into a global pandemic. Whilst often considered a respiratory disease, a large proportion of COVID-19 patients report neurological symptoms, and there is accumulating evidence for neural damage in some individuals, with recent studies suggesting loss of gray matter in multiple regions, particularly in the left hemisphere. There are a number of mechanisms by which COVID-19 infection may lead to neurological symptoms and structural and functional changes in the brain, and it is reasonable to expect that many of these may translate into cognitive problems. Indeed, cognitive problems are one of the most commonly reported symptoms in those suffering from “Long COVID”—the chronic illness following COVID-19 infection that affects between 10–25% of sufferers. The COVID and Cognition Study is a part cross-sectional, part longitudinal, study documenting and aiming to understand the cognitive problems in Long COVID. In this first paper from the study, we document the characteristics of our sample of 181 individuals who had suffered COVID-19 infection, and 185 who had not. We explore which factors may be predictive of ongoing symptoms and their severity, as well as conducting an in-depth analysis of symptom profiles. Finally, we explore which factors predict the presence and severity of cognitive symptoms, both throughout the ongoing illness and at the time of testing. The main finding from this first analysis is that that severity of initial illness is a significant predictor of the presence and severity of ongoing symptoms, and that some symptoms during the acute illness—particularly limb weakness—may be more common in those that have more severe ongoing symptoms. Symptom profiles can be well described in terms of 5 or 6 factors, reflecting the variety of this highly heterogenous condition suffered by the individual. Specifically, we found that neurological and fatigue symptoms during the initial illness, and that neurological, gastro-intestinal, and cardiopulmonary symptoms during the ongoing illness, predicted experience of cognitive symptoms.

## 1 Introduction

 Since its first emergence in December 2019, coronavirus disease 2019 (COVID-19), which is caused by severe acute respiratory syndrome coronavirus 2 (SARS-CoV-2), has evolved into a global pandemic (WHO 2020). As of July 27, 2021, there have been over 200 million confirmed cases causing more than 4 million deaths across 237 countries worldwide (WHO 2021).

Manifestations of COVID-19 infection vary in severity ranging from asymptomatic to fatal. In the acute stage, symptomatic patients—at least in the early variants—typically experience respiratory difficulties that can result in hospitalization and require assisted ventilation (Baj et al. 2020; Heneka et al. 2020; Jain 2020). While COVID-19 is primarily associated with respiratory and pulmonary challenge, 35% of patients report neurological symptoms including headache, dizziness, myalgia or loss of taste and smell (e.g., Mao et al. 2020). In severe illness, neurological symptoms can be seen in 50–85% of patients (e.g., Pryce-Roberts, Talaei, and Robertson 2020; Romero-Sánchez et al. 2020). Indeed, alteration in taste or smell (anosmia/dysgeusia) is often the first clinical symptom (Mao et al. 2020; Romero-Sánchez et al. 2020) and is reported in over 80% of cases (e.g., Lechien et al. 2020). Furthermore, this symptom regularly persists beyond resolution of respiratory illness (Lechien et al. 2020). It has been argued that many of the non-neurological symptoms characteristic of COVID-19 infection may in fact reflect viral invasion of brain stem and hypothalamus, leading to dysregulation of basic autonomic functions such as respiration, cardiovascular function, thermoregulation and glycemia (e.g., Mussa, Srivastava, and Verberne 2021). The details of the symptom pattern may vary with patient age, infecting variant or both, an area of active investigation at the time of writing.

Accumulating evidence suggests that many patients suffering from severe illness show evidence of neural damage. A study of COVID-19 intensive care (ICU) patients found that those with neurological symptoms demonstrated gray matter abnormality (Kandemirli et al. 2020) or other non-specific neural abnormalities (Helms et al. 2020). Post-mortem studies of patients who have died of COVID-19 show evidence for ischemic lesions and indications of neuro-inflammation (Matschke et al. 2020). Abnormalities—including hemorrhagic lesions— have been identified in the orbitofrontal cortex (Le Guennec et al. 2020), medial temporal lobe and hippocampus (Moriguchi et al. 2020; Poyiadji et al. 2020), bilateral thalami, and subinsular regions (Poyiadji et al. 2020). There is also evidence for unusual neural activity: Nearly 90% of electroencephalography (EEG) studies performed in COVID-19 patients revealed epileptiform discharges mostly within the frontal lobes (Galanopoulou et al. 2020). A recent study using the UK Biobank cohort conducted structural and functional brain scans before and after infection with COVID-19 on 394 patients compared with 388 matched controls who had not experienced COVID-19 infection (Douaud et al. 2021). These participants were all over 45 and had 3 years between scans. In a region of interest analysis (based on hypothesized viral spread from the olfactory bulb), the authors identified significant loss of gray matter in the parahippocampal gyrus, lateral orbitofrontal cortex and insula, notably concentrated in the left hemisphere. Exploratory analysis of the entire cortical surface supported this pattern. While only a small subset of this sample (*N* = 15) had experienced severe disease and hospitalization, there was some evidence to suggest that these individuals had more severe gray matter loss, with potentially greater effects in the left cingulate cortex, and right amygdala and hippocampus.

Given the evidence for widespread neural symptoms and demonstrable neural damage, it could be expected that COVID-19 infection would be associated with cognitive deficits. The form that these deficits take should in principle be relatable to the place, extent and nature of the underlying neuropathology (Bougakov, Podell, and Goldberg 2021). There are a number of postulated mechanisms linking COVID-19 infection with neurological problems. It has been hypothesized, based on the behavior of previous SARS viruses, that SARS-CoV-2 can attack the brain directly perhaps via the olfactory nerve (Lechien et al. 2020; Politi, Salsano, and Grimaldi 2020) causing encephalitis. Severe hypoxia from respiratory failure or distress can also induce hypoxic/anoxic-related encephalopathy (Guo et al. 2020). There is considerable evidence that COVID-19 is associated with abnormal blood coagulation, which can increase risk of acute ischemic and hemorrhagic cerebrovascular events (CVAs) (Beyrouti et al. 2020; Kubánková et al. 2021; Li et al. 2020; Wang et al. 2020) leading to more lasting brain lesions. These brain changes would be unlikely to be present uniquely as cognitive deficits, but would be associated with a range of related symptoms. Some of these symptoms may be neurological (e.g., disorientation, headache, numbness) while others may reflect systemic/multi-system involvement (reflecting the symptom profile of chronic inflammatory or autoimmune diseases for example). It may therefore be possible to gain information as to the mechanism of neurological involvement via investigation of symptomatology.

It has been suggested that the action of the immune system itself can be deleterious. Dysfunctional or excessive immune response to infection can take a number of forms, each associated with negative impact on neural systems. For example, excessive cytokine release (“cytokine storm”) can result in hemorrhagic encephalopathy (Das, Mukherjee, and Ghosh 2020; Poyiadji et al. 2020). Alternatively, immune-mediated peripheral neuropathy (e.g., Guillain-Barre syndrome) can occur, characterized by limb weakness, loss of deep tendon reflexes with sensory abnormalities (Alberti et al. 2020; Whittaker, Anson, and Harky 2020; Zhao et al. 2020).

Before the COVID-19 pandemic, there was already considerable evidence that inflammation— particularly chronic inflammation—is associated with neural and cognitive dysfunction. In rodent models, it has been demonstrated that inflammation following infections is associated with disrupted neurogenesis in the hippocampus, both via reduced differentiation and survival of new cells (Ekdahl et al. 2003; Monje, Toda, and Palmer 2003) and disrupted integration of new cells into existing hippocampal networks (Belarbi et al. 2012; Jakubs et al. 2008). Systemic inflammation leads to significant reductions in brain-derived neurotrophic factor (BDNF) in hippocampus and cortical regions (e.g., Chapman et al. 2012; Guan and Fang 2006; Lapchak, Araujo, and Hefti 1993; Schnydrig et al. 2007) and multiple inflammatory cytokines are linked with evidence of cognitive impairments (e.g., IL-1β: Beilharz, Maniam, and Morris 2014; Beilharz et al. 2018; Che et al. 2018; Mirzaei et al. 2018; Thirumangalakudi et al. 2008; TNF-α: Almeida-Suhett et al. 2017; Beilharz et al. 2014; Thirumangalakudi et al. 2008). These findings are broadly reflected in human studies, wherein circulating cytokines have been associated with reduced episodic memory and greater number of Alzheimer’s symptoms (e.g., Kheirouri and Alizadeh 2019) and chronic neuroinflammation has been heavily implicated in the pathophysiology of neurodegenerative diseases (Bossù et al. 2020; Chen, Zhang, and Huang 2016; McGeer and McGeer 2010; Zotova et al. 2010). Given the volume of reports of excessive immune response to COVID-19 infection (Mehta et al. 2020; Tay et al. 2020), some neural and cognitive disruption is therefore unsurprising, but the nature, timing and extent become important areas for research.

There is some early evidence linking neural changes following COVID-19 and cognitive deficits. Hosp and colleagues (2021) studied 29 patients (average age 65) with no history of cognitive impairment or neurodegeneration, and presenting at least one new neurological symptom since COVID-19 infection. Positron emission tomography (PET) analysis revealed pathological results with predominant frontoparietal hypometabolism, correlating to lower scores on the Montreal Cognitive Assessment (MoCA) and extended neuropsychological testing. In particular, COVID-19 patients showed deficits in tests of verbal memory and executive functions (Hosp et al. 2021).

The UK Office for National Statistics (ONS 2021) has estimated that around 21% of those suffering COVID-19 infection still have symptoms at 5 weeks, and that 10% still have symptoms at 12 weeks from onset. These figures may not tell the full story, being based on a list of 12 symptoms which does not include neurological or cognitive manifestations (e.g., Alwan and Johnson 2021; Ziauddeen et al. 2021). Other calculations suggest that around 1 in 3 non-hospitalized COVID-19 patients have symptoms after 2–6 weeks from disease onset (Nehme et al. 2021; Sudre et al. 2020; Tenforde et al. 2020) and that 11–24% still have persisting symptoms 3 months after disease onset (Cirulli et al. 2020; Ding et al. 2020). A community-based study reported that around 38% symptomatic people experienced at least one symptom lasting 12 weeks or more from onset and around 15% experienced three or more symptoms (Whitaker et al. 2021). Ongoing symptoms seem to occur regardless of the severity of the initial infection, with even asymptomatic patients sometimes going on to develop secondary illness (FAIR Health 2021; Nehme et al. 2021), however initial severity may impact severity of ongoing issues (e.g., Whitaker et al. 2021).

The National Institute for Health and Care Excellence (NICE) guidelines describe “post-COVID-19 syndrome” as “*Signs or symptoms that develop during or after infection consistent with COVID-19, continue for more than 12 weeks and are not explained by an alternative diagnosis”* (NICE 2020). One difficulty with this definition is that the “signs or symptoms” that qualify for the diagnosis are not specified (e.g., Alwan and Johnson 2021; Ziauddeen et al. 2021) thus many sufferers could go uncounted and unrecognized clinically, or conversely over-liberal inclusion may lead to overcounting. The patient-created term “Long COVID” has increasingly been used as an umbrella term to describe the highly heterogenous condition experienced by many people following COVID-19 infection (Callard and Perego 2021).

Emerging evidence suggests that Long COVID is a debilitating multisystem illness and there have been some attempts to characterize “phenotypes”. An online survey involved in 2550 non-hospitalized participants detected two clusters within both acute and ongoing symptoms. Acute symptoms showed a majority cluster with cardiopulmonary symptoms predominant, and a minority cluster with multi-system symptoms. Similarly, ongoing symptoms were clustered into a majority cluster with cardiopulmonary, cognitive symptoms and exhaustion, and a minority cluster with multisystem symptoms. Those with more related symptoms in the acute major cluster were more likely to move into ongoing multisystem cluster, and this movement can be predicted by gender and age, with higher risk in women, those younger than 60, and those that took less rest during the initial illness (Ziauddeen et al. 2021).

“Long COVID” research has repeatedly identified cognitive dysfunction as one of the most common persistent symptoms (after fatigue), occurring in around 70% of patients (Bliddal et al. 2021; Cirulli et al. 2020; Davis et al. 2021; Ziauddeen et al. 2021). Indeed, brain fog and difficulty concentrating are more common than cough is at many points in the Long COVID time course (Assaf et al. 2020). Ziauddeen and colleagues report nearly 40% of participants endorsing at least one cognitive symptom during the initial two weeks of illness, with this persisting in the long term. However around 30% of participants also reported developing cognitive symptoms—particularly brain fog and memory problems— later (Ziauddeen et al. 2021). Indeed, Davis and colleagues (2021) demonstrate that brain fog, memory problems and speech and language problems were more commonly reported at week 8 and beyond than they were during initial infection. Furthermore, strenuous cognitive activity was found to be one of the most common triggers leading to relapse/exacerbation of existing symptoms (Davis et al. 2021; Ziauddeen et al. 2021). Crucially, 86% of participants indicated that cognitive dysfunction and/or memory impairment was impacting their ability to work, with nearly 30% reporting being “severely unable to work” and only 27% working as many hours as they had pre-COVID-19 (Davis et al. 2021). These figures suggest that the cognitive sequelae of COVID-19 have the potential for long-term consequences not just for individuals but also—given the prevalence of Long COVID—for the economy and wider society.

Here we report on the first stage of a mixed cross-sectional/longitudinal investigation—The COVID and Cognition Study (COVCOG)—aimed at understanding cognition in post-acute COVID-19.

The study consists of a baseline assessment of characteristics and cognition in samples of individuals who have or have not experienced COVID-19 infection. Both groups complete a range of cognitive tasks and are then followed up at regular intervals. Using the online assessment platform Gorilla (www.gorilla.sc), we set out to bring together information about symptom profiles both during and following initial infection and detailed analysis of cognitive performance across a range of domains including memory, language and executive function. The current paper reports on the characteristics of the cohort, including (in those that have experienced COVID-19) a detailed investigation of symptom profiles. Individuals who had experienced COVID-19 infection reported on a large selection of symptoms for three time periods: the initial infection (first 3 weeks), the time “since then”, and “currently” (the time of test and 1–2 days preceding). This allows investigation of symptoms during initial illness that may be predictive of ongoing symptoms, as well as exploring the nature of those ongoing symptoms themselves. These symptom analyses provide stratifiers and covariates for following papers that will report on cognitive test performance at the baseline, and track changes in symptoms and cognition through analysis of follow-up assessments.

The aims of this initial analysis are three-fold: First, to establish similarity in our small sample relative to other, larger and more comprehensive, studies of Long COVID. Due to the intensive performance focus of the current investigation, we are limited to a smaller sample than is feasible in an epidemiological cohort. However, if our sample is, at broadly, similar to those in other larger studies, then it may be possible to generalize our findings more widely.

Second, we aim to contribute to the understanding of phenotypes of Long COVID by using a rigorous factor analytic approach to identify groups of symptoms that tend to co-occur. In a highly heterogenous condition, in which up to 200 symptoms have been suggested (Davis et al. 2021), reduction of dimensionality is essential to allow meaningful associations to be drawn between experienced symptoms and relevant outcomes. Systematic symptomatology requires not merely judging the labels applied to factors to be qualitatively interpretable (i.e., show a coherent item loading pattern) but also comparing alternative structures quantitatively in fit, parsimony and agreement with other evidence. Later studies can then benefit by proceeding in a way more characteristic of a test development programme; better supporting the weaker factors by addition of appropriate items. Thus, this first study need not finalize the adopted number of factors, but can assist in the resolution of that question in subsequent studies, and so support new topic findings via evolving methodology.

In an application of this second aim, a third objective is to use the symptom factors extracted to investigate predictors of self-reported cognitive deficits. If it is possible to identify groups of symptoms during either the acute-phase, or during the post-acute (“ongoing”) illness that predict cognitive problems, this may aid in the identification of patients that are at risk of developing cognitive deficits. While this study is not able to identify a specific mechanism (as this requires types of tests and analysis not feasible to conduct online) it may be able to lay the groundwork with sufficient breadth and detail to inform future mechanistic investigation.

Due to the novel character of both the virus and the subsequent ongoing illness at the time of study creation, this study was designed not to test specific hypotheses but to map the terrain, generating hypotheses for future, more targeted investigation. However, we are able to offer some tentative hypotheses.

H1: Sample Characteristics: First, we predict that our sample will, in general, reflect patterns already demonstrated in other studies of Long COVID—including age and gender effects, prevalence of cognitive symptoms and degree of impact on everyday functioning.

H2: Symptom Profiles: We predict that the factors emerging from the symptom analysis may indicate Long COVID “phenotypes” which may, through future studies, be directly linked to disease profiles and mechanisms.

H3: Cognitive Symptoms: Finally, we predict that some of these symptom factors— particularly those with incorporating neurological symptoms—will be predictive of self-reported cognitive deficits.

## 2 Methods

### 2.1 Participants

A total of 421 participants aged 18 and over were recruited through word of mouth, student societies and online/social media platforms such as the Facebook *Long COVID Support Group* (over 40K members). Of these, 163 participants were recruited through the *Prolific* recruitment site, targeting participants with demographic profiles underrepresented in our sample. Specifically, recruitment through *Prolific* was limited to those with low socioeconomic status and levels of education below a bachelor’s degree. As the study was conducted in English, participants were recruited from majority English speaking countries (the UK, Ireland, US, Canada, Australia, New Zealand, or South Africa). Informed consent to use of anonymized data was obtained prior to starting.

Data collection for this stage of the study took place between October 2020 and March 2021, and recorded data on infections that occurred between March 2020 and February 2021. As such, all participants with experience of COVID-19 infection were likely to have been infected with either Wild-Type or Alpha-variant SARS-CoV-2, as the later-emerging Delta variant was not common in the study countries at that time. Study recruitment started before the roll out of vaccinations, thus we do not have confirmed vaccination status for all participants. Once vaccination became available, the questionnaire was revised to ask about vaccination status. Of the 33 participants who were tested after this point, 11 (2 in the No COVID group, 9 in the COVID group) reported being vaccinated. Among them, 8 received the first dose and 3 already received the second dose. The majority (over 80%) had the vaccine within the last 7 days to last month. All received Pfizer (BNT162b2) except 1 (COVID group) who received AstraZeneca (AZD1222).

### 2.2 Procedure

The study was reviewed and a favorable ethics opinion was granted by University of Cambridge Department of Psychology ethics committee (PRE.2020.106, 8/9/2020). This is a mixed cross-sectional/longitudinal online study conducted using the Gorilla platform (www.gorilla.sc). The results reported here are for the baseline session of the study only. The baseline session consisted of a questionnaire covering demographics, previous health and experience of COVID-19, followed by a series of cognitive tests. The cognitive tests will be reported in a following publication.

Participants answered questions relating to their age, sex, education level, country of permanent residence, ethnicity, and profession. They were then asked a series of questions relating to their medical history, and health-related behaviors (such as smoking and exercise).

Following this, they were asked for details of their experience of COVID-19. Because many of the participants in this study contracted COVID-19 before confirmatory testing of infection state was widely available, both those with and without test confirmation were included in the “COVID” group. Those that didn’t think they had had COVID-19 but had experienced an illness that *could* have been COVID-19 were assigned an “unknown” infection status. Those that confirmed that they had not had COVID-19, nor any illness that might have been COVID-19, were included in the “No COVID” group.

The procedure for progression through the baseline session is detailed in Figure 1. Participants in the “No COVID” Group proceeded directly to cognitive tests. Participants in the “COVID” group indicated the number of weeks since infection on a drop-down menu. Those that reported being within the first 3 weeks of infection proceeded straight to debriefing and were followed up 2 weeks later, once the initial infection was passed. Apart from this delay, they proceeded with the experiment in the same way as the rest of the COVID group.

**Figure 1.**
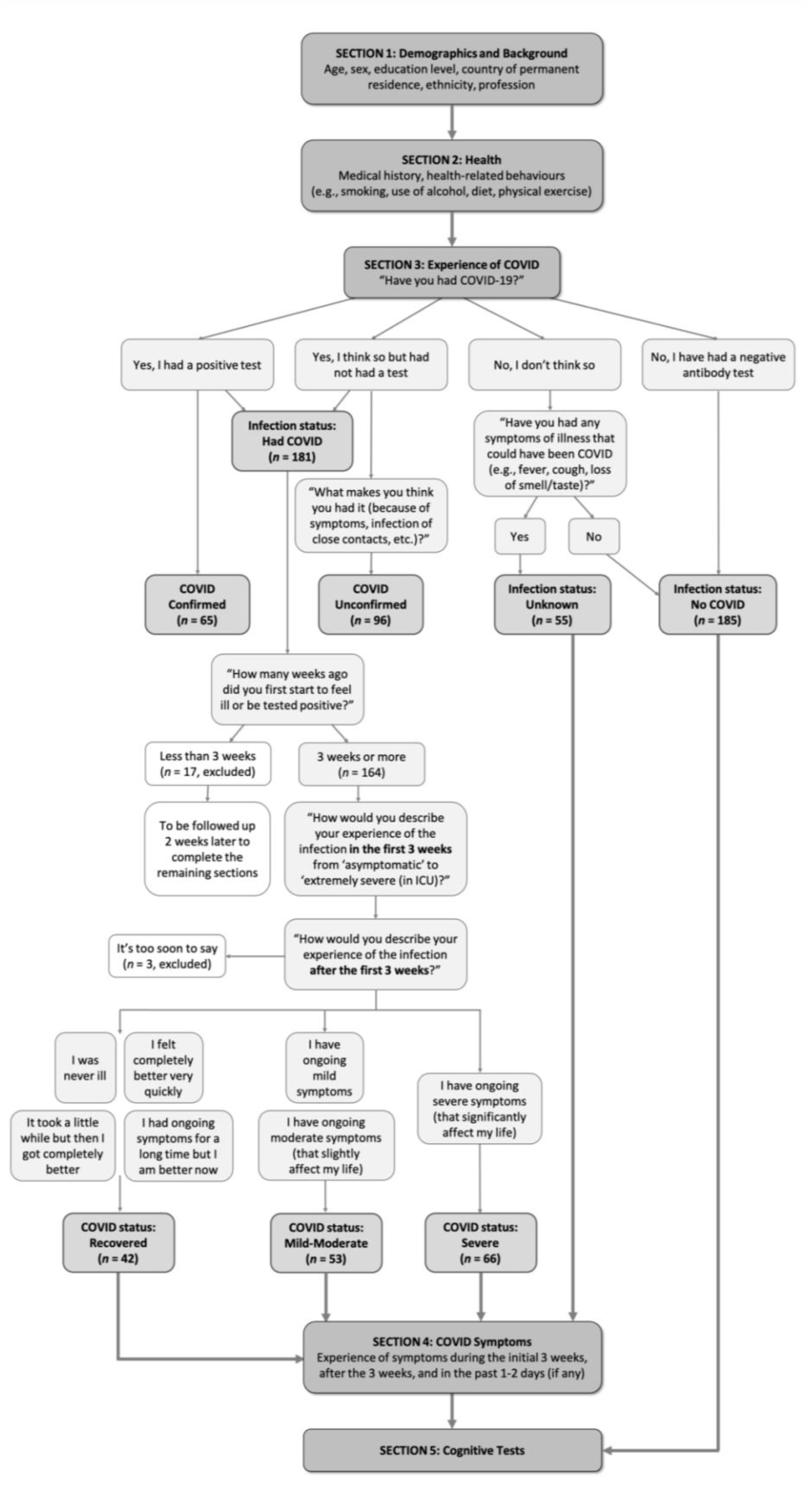
Study Procedural flow.

Participants then answered questions on the severity of the initial illness and whether they were experiencing ongoing symptoms. Finally, Participants were asked to give details on a large number of individual symptoms during three time periods: When reporting on initial symptoms, participants gave an indication of severity on a scale of 1–3. When reporting symptoms over the period “since then”, they reported on both severity and regularity of symptoms on a scale of 1–5. When reporting on symptoms in the past 1–2 days, they reported the presence or absence of the symptoms.

### 2.3 Data Processing and Analysis

Analyses were conducted using IBM SPSS Statistics for Windows, Version 23.0. We describe quantitative variables using means and standard deviations, and numbers and percentages for qualitative variables. Sidak’s correction for multiple comparisons was employed. All *p* values are reported uncorrected, and the Sidak-corrected alpha is quoted where appropriate.

We investigated differences in the first group of variables: sociodemographic, medical history, and health behaviors, concerning two COVID group classifications. First dividing the sample into two groups (COVID/No COVID), second subdividing the COVID group by symptom longevity and severity (Recovered, Ongoing mild infection, and Ongoing severe infection). Where parametric analysis was not appropriate, we employed the Pearson’s chi-square (*χ^2^*) test for categorical variables and the Mann-Whitney and Kruskal-Wallis test for continuous variables depending on the number of COVID groups. To explore what variables were associated with infection or ongoing symptoms we employed various independent multinomial logistic regression models with backward deletion of variables *p* > .05. To investigate differences between groups (COVID/No COVID; Recovered / Ongoing mild / Ongoing severe) we employed independent t-test/Mann-Whitney and ANOVA/Kruskal-Wallis.

## 3 Results

### 3.1 Sample Characteristics

#### 3.1.1 No COVID (*NC*: *n* = 185) vs COVID (*C*: *n* = 181)

Distributions of demographics including sex, age, education level, country and ethnicity of the two groups (*NC*/*C*) are shown in Table 1. The majority of participants were from the United Kingdom and were of White (Northern European) ethnicity (over 70% in both groups). Pearson’s chi-square tests showed that the groups did not significantly differ in sex, but differed in age (*χ^2^*[5] = 19.08, *p* = .002, *V* = .228) and level of education (*χ^2^*[5] = 56.86, *p* < .001, *V* = .394), with the COVID group tending to fall into the older age ranges and higher education level more than the No COVID group.

**Table 1.**
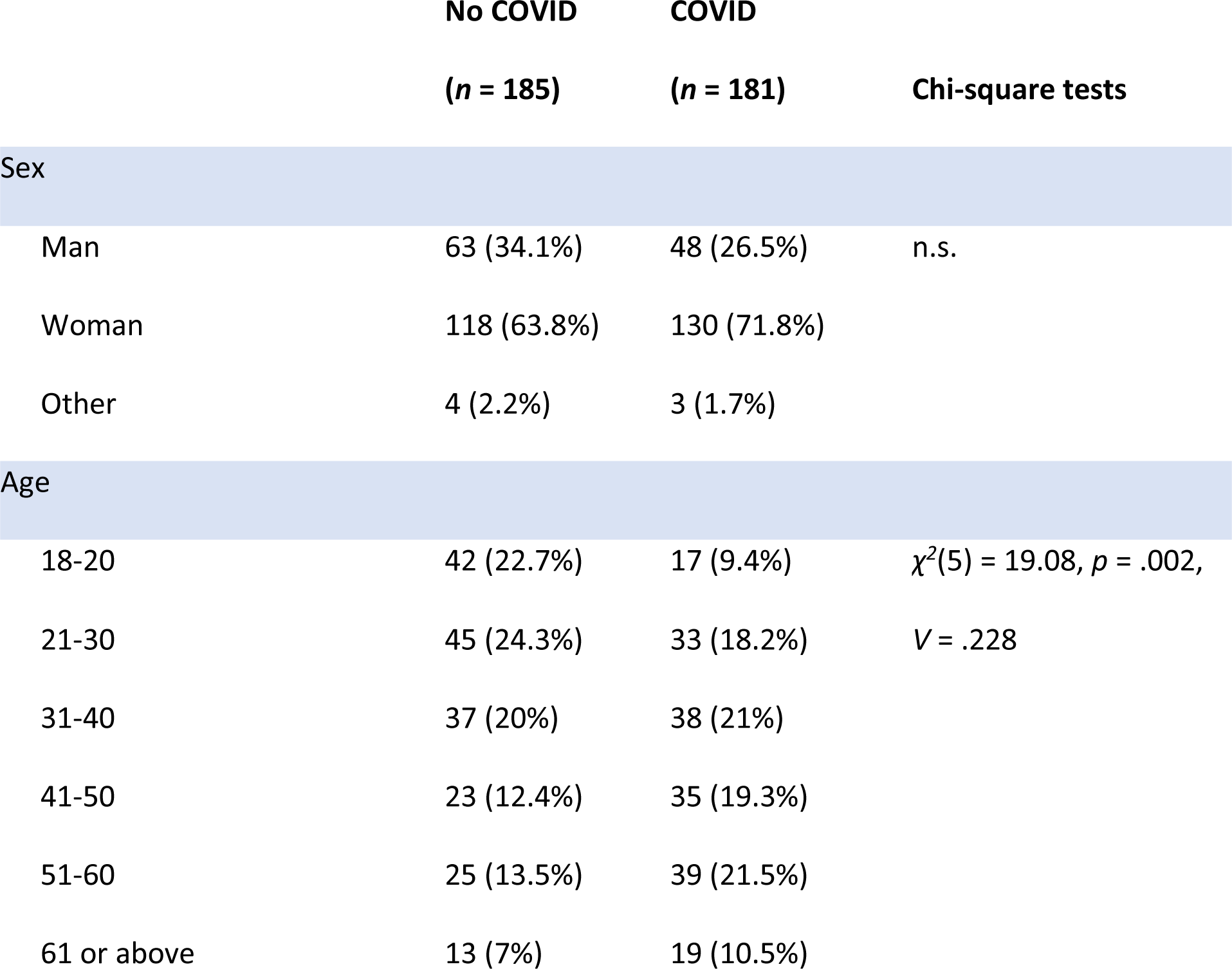
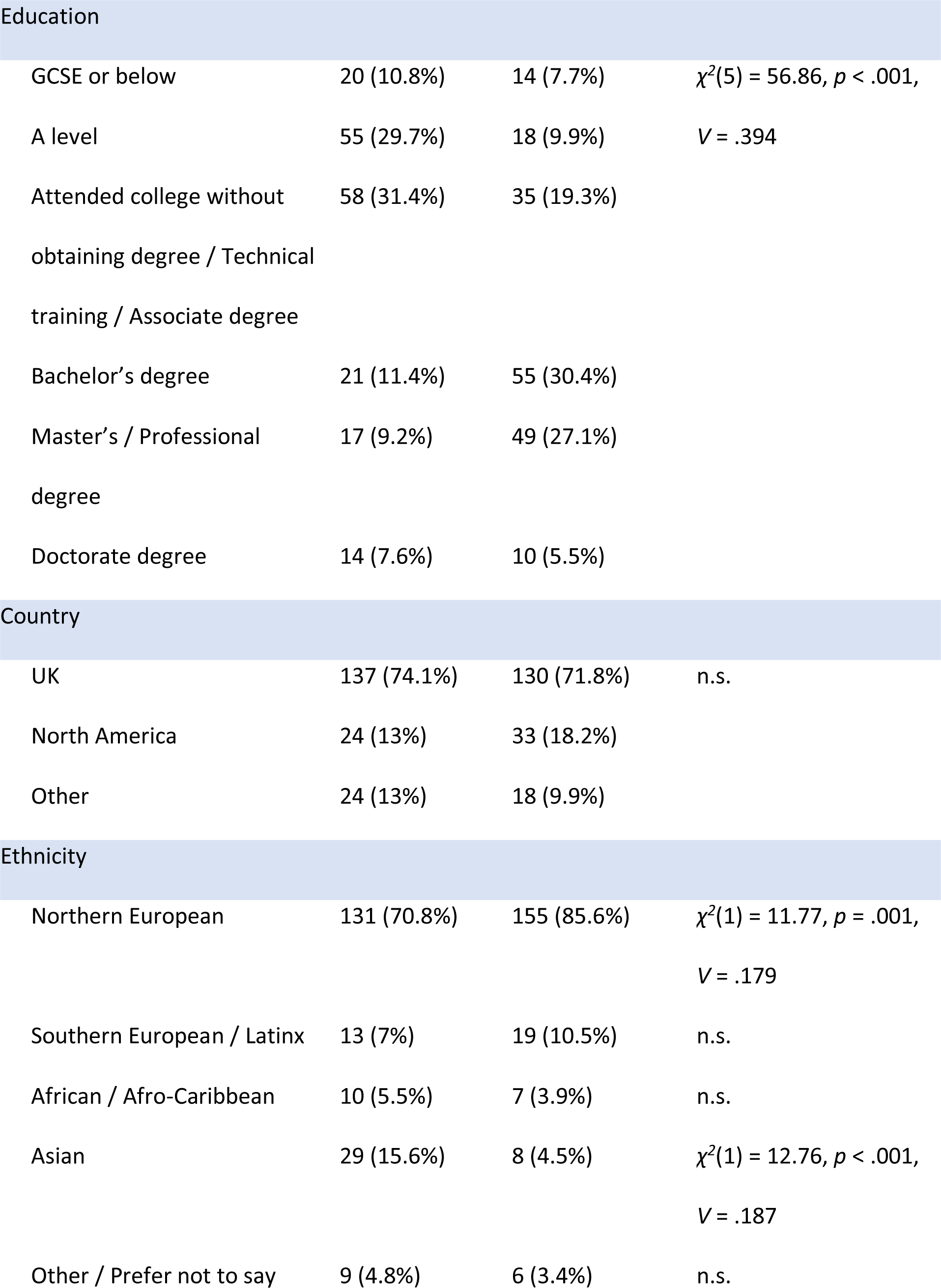
Distribution of demographics in No COVID and COVID groups.

**Table 2.**
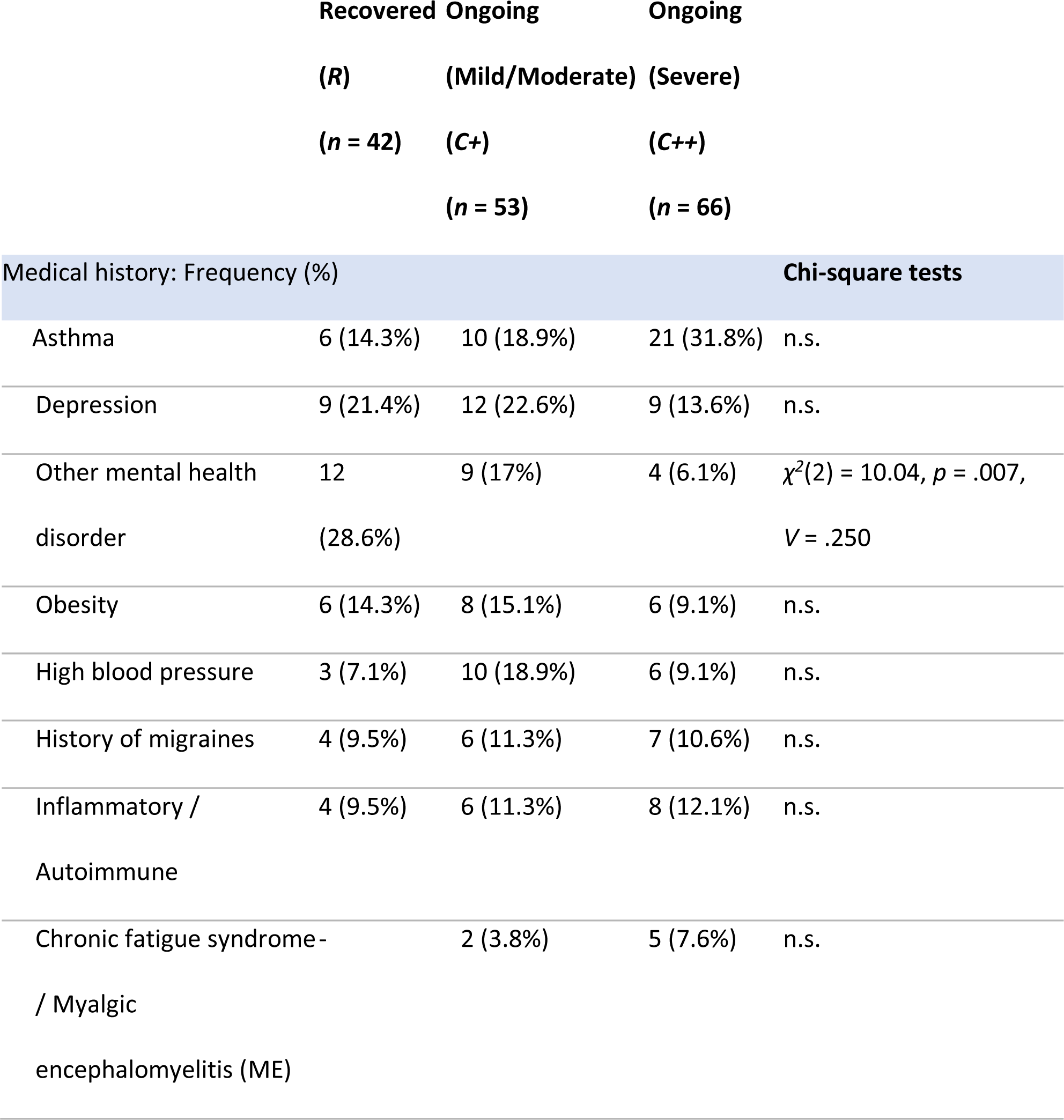
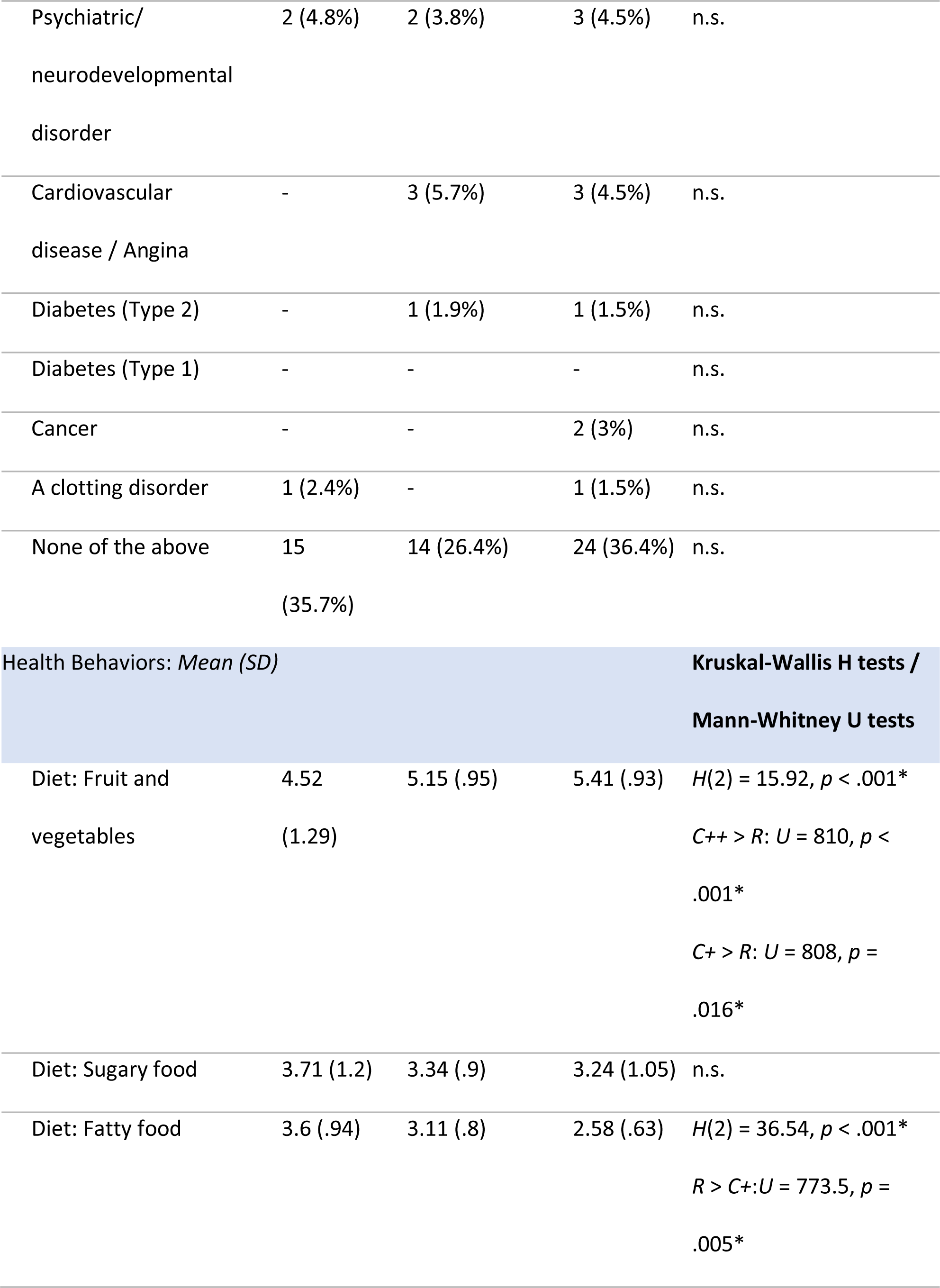
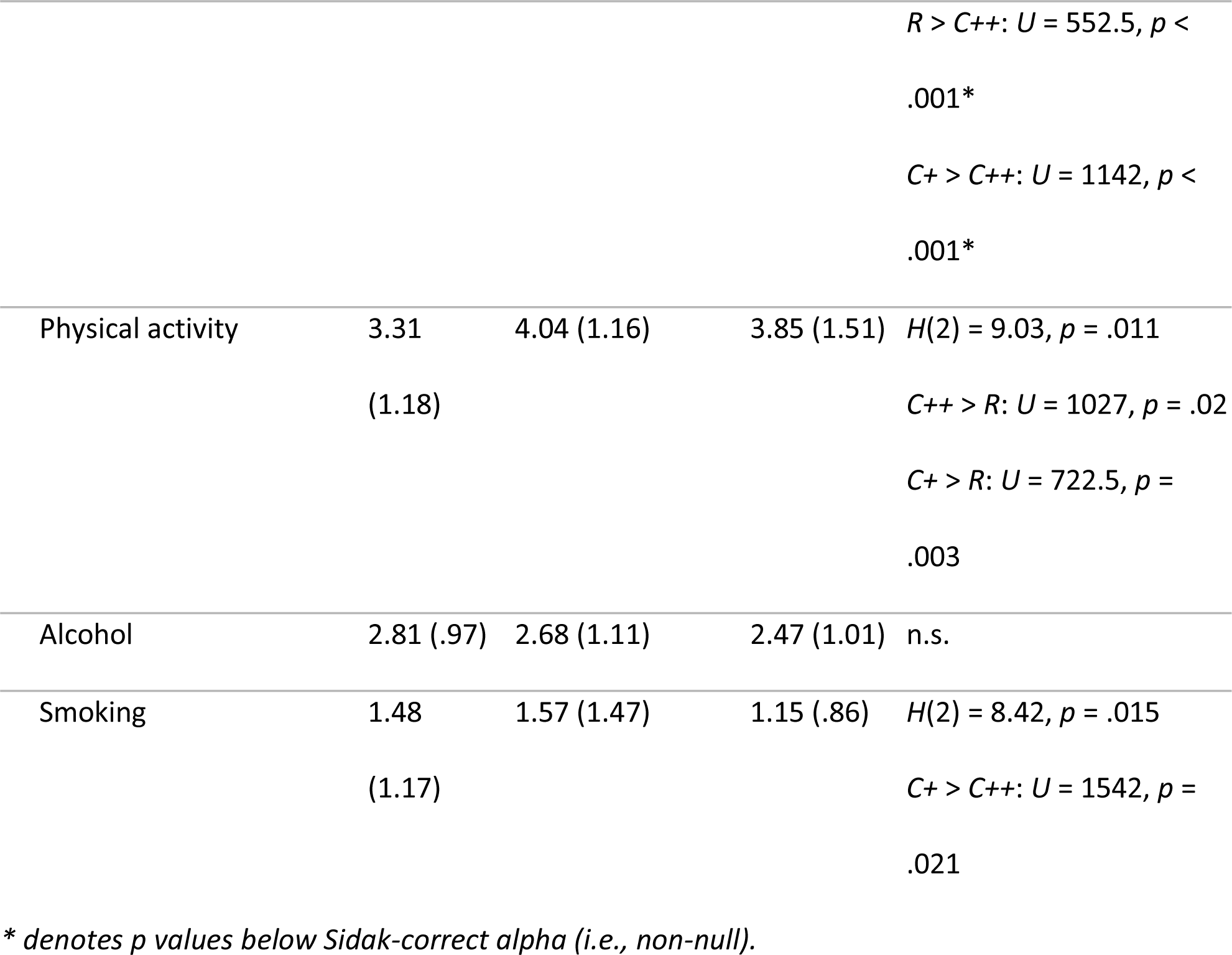
Distribution of medical history and health behaviors (1 = *Never* – 6 = *Several times daily*; higher scores indicating higher frequency) in COVID subgroups: Recovered (*R*), Ongoing (Mild/Moderate (*C+*)) and Ongoing (Severe (*C++*)).

#### 3.1.2 Employment

Supplementary Table 1 shows the distributions of pre-pandemic profession and employment status. To adjust for multiple comparisons, Sidak corrections were applied and alpha levels were adjusted to .003 for profession and .007 for employment status. The COVID group had significantly more people working in healthcare (*χ^2^*[1] = 12.77, *p* < .001, *V* = .187) and engaging in full-time work before the pandemic (*χ^2^*[1] = 21.19, *p* < .001, *V* = .241). In contrast, the No COVID group were more likely not to be in paid work (Profession “Not in paid work” *χ^2^*[1] = 27.72, *p* < .001, *V* = .275; Employment status “Not Working” *χ^2^*[1] = 13.18, *p* < .001, *V* = .190), and they were more likely to be students (*χ^2^*[1] = 8.91, *p* = .003, *V* = .156).

#### 3.1.3 Health and Medical History

Supplementary Table 2 compares medical history and health behaviors across the COVID and No COVID groups, which may be informative as to vulnerabilities. Sidak correction adjusted the alpha level to .003 for medical history and .008 for health behaviors. Pearson’s chi-square tests showed that inflammatory or autoimmune diseases (*χ^2^*[1] = 9.81, *p* = .002, *V* = .164) were found more commonly in the COVID group than the No COVID group. Mann-Whitney U tests showed that the COVID group consumed more fruit and vegetables (*U* = 13525, *p* = .001) and had higher level of physical activity (*U* = 13752, *p* = .002) than the No COVID group, while the No COVID group consumed sugary (*U* = 14168.5, *p* = .008) food more than the COVID group. ANOVA showed that the COVID group (*M* = 26.71, *SD* = 7.26) had higher body mass index (BMI) than the No COVID group (*M* = 25.15, *SD* = 5.64), *F*(1,361) = 5.24, *p* = .023. However this effect was not significant after controlling for sex, age, education and country (*F*(1,357) = 1.57, *p* = .211).

### 3.2 Characteristics of Those Experiencing Ongoing Symptoms

To understand the potential association between the progression of COVID-19 and various potential risk factors at baseline, including demographics, medical history and health behaviors, and the severity of initial illness and initial symptoms, we further divided the COVID group into three duration subgroups: (i) those who, at the time of test, had recovered from COVID-19 (“Recovered group”, *R*; *n* = 42), (ii) those who continued to experience mild or moderate ongoing symptoms (“Ongoing (Mild/<u>M</u>oderate) group”, *C+*; *n* = 53), and (iii) those who experienced severe ongoing symptoms (“Ongoing (Severe) group”, *C++*; *n* = 66). Those who were still at their first 3 weeks of COVID-19 infection (*n* = 17) or those who reported “it is too soon” to comment on their ongoing symptoms (*n* = 3) were not included in the following analyses. Participants in all groups ranged between 3 and 31+ weeks since symptom-onset, and a majority (81.5%) of those with ongoing symptoms reporting after more than 6 months since infection.

Figure 2 shows the distribution of demographic variables across the COVID-19 duration subgroups (further details available in Supplementary Table 3). In each, more than half of the participants were from the United Kingdom (54.8–92.4%) and were of White (Northern European) ethnicity (69–93.9%). Pearson’s chi-square tests suggested that age (*χ^2^*[10] = 53.41, *p* < .001, *V* = .407) and education level (*χ^2^*[10] = 20.03, *p* = .029, *V* = .249), but not sex, significantly differed between subgroups. In terms of age, the *R* subgroup tended to fall more in the younger age ranges (see Figure 2a). In terms of education level, the *R* subgroup tended to have lower education level (GCSE or below and A level), but the *C++* (Severe) subgroup clustered more in higher education level (bachelor’s degree) (see Figure 2b). The subgroups also differed in the time elapsed since infection at the time of completing the study (*χ^2^*[6] = 19.64, *p* = .003, *V* = .247). The *R* subgroup were more likely to be in their first 10 weeks of infection, while the *C++* (Severe) subgroup were more likely to be at their 31 weeks or above (Figure 1c).

**Figure 2.**
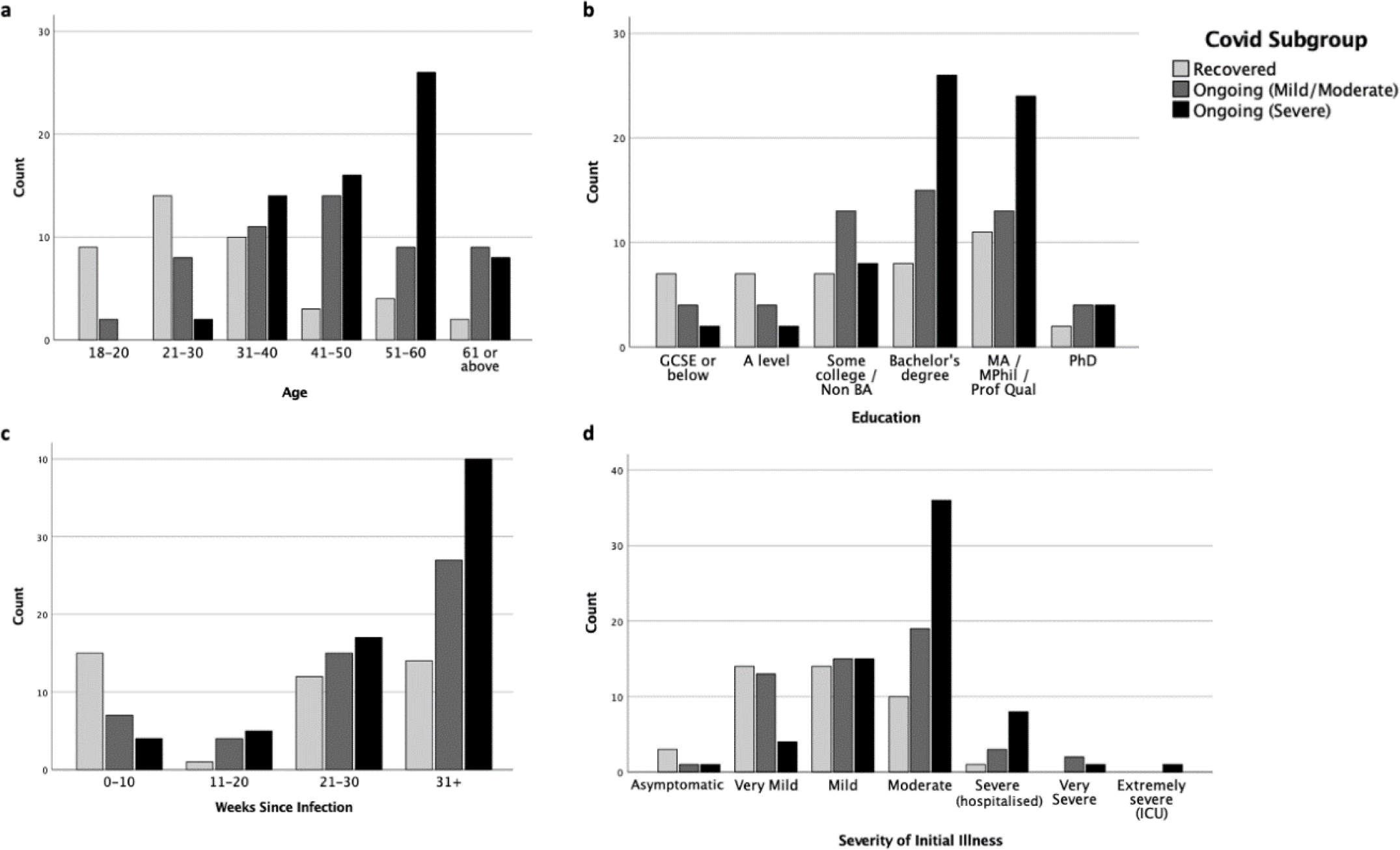
Distributions of a) age, b) education level, c) weeks since infection, and d) severity of initial illness in Recovered, Ongoing (Mild/Moderate) and Ongoing (Severe) subgroups.

A multinomial logistic regression indicated that only age, but not sex or education, significantly predicted COVID-19 progression (*χ^2^*[10] = 43.6, *p* < .001). People in the age ranges of 18–20 years and 21–30 years were more likely to recover from COVID-19 than to progress into mild/moderate (*p*s = .02–.03) or severe (*p* = .002) ongoing symptoms.

We examined whether medical history and health behaviors were associated with different degrees of ongoing symptoms. Tables 5 shows the descriptive statistics of these factors in *R*, *C+* and *C++* subgroups for medical history and pre-pandemic health behaviors. None of the listed health conditions were significantly related to ongoing symptom severity (against Sidak *α* = .003). There were, however, significant group differences (Sidak *α* = .008) in fruit and vegetables consumption (*H*[2] = 15.92, *p* < .001) and fatty food consumption (*H*[2] = 36.54, *p* < .001). Both ongoing symptom subgroups ate more fruit and vegetables (*C++*: *U* = 810, *p* < .001; *C+*: *U* = 808, *p* = .016) and less fatty food (*C+*: *U* = 773.5, *p* = .005; *C++*: *U* = 552.5, *p* < .001) than the *R* subgroup. The *C+* (Mild/Moderate) subgroup also consumed more fatty food than the *C++* (Severe) subgroup (*U* = 1142, *p* < .001). The subgroups did not significantly differ in BMI (*F*[2,157] = .085, *p* = .919).

**Table 3.**
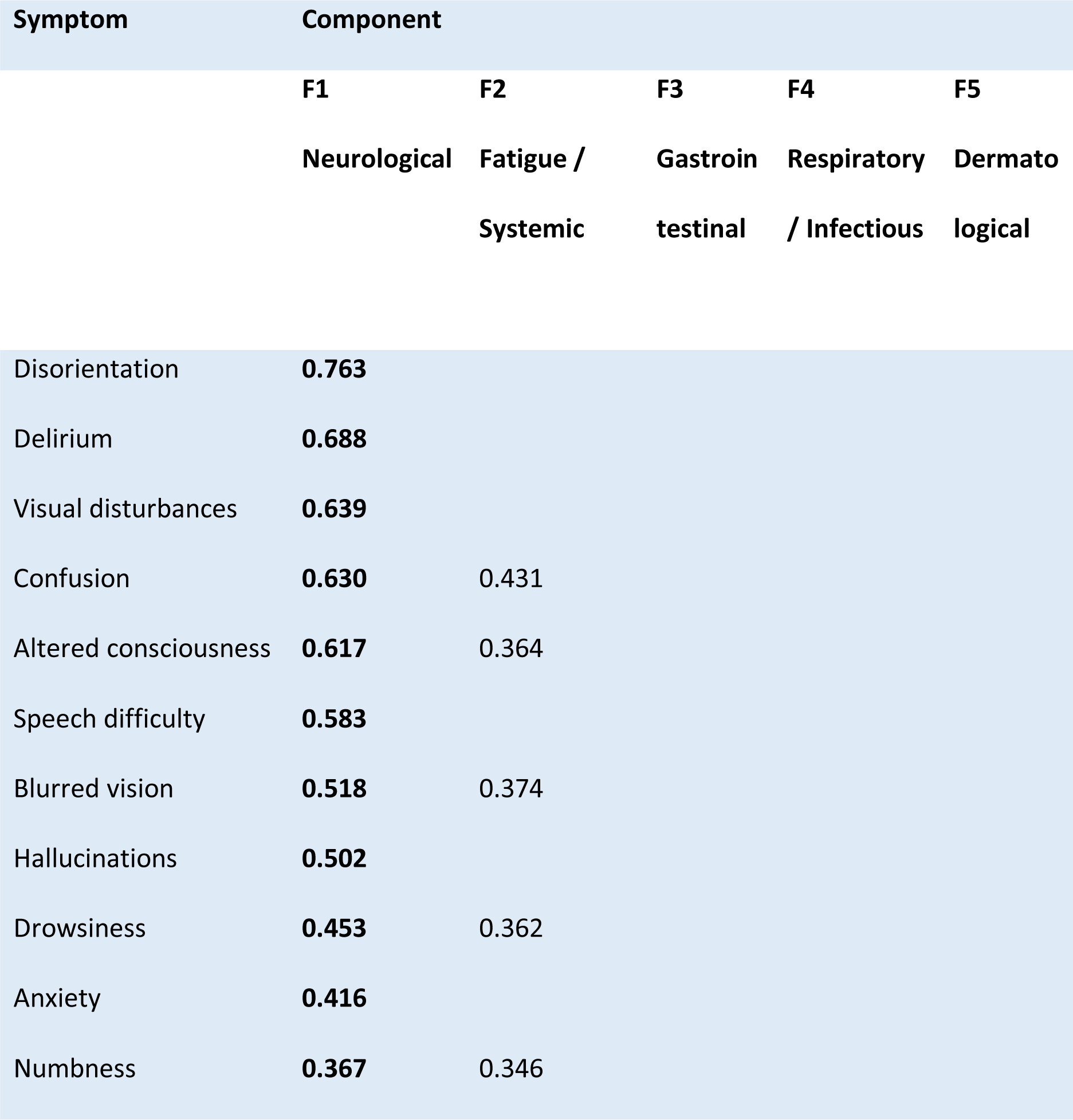
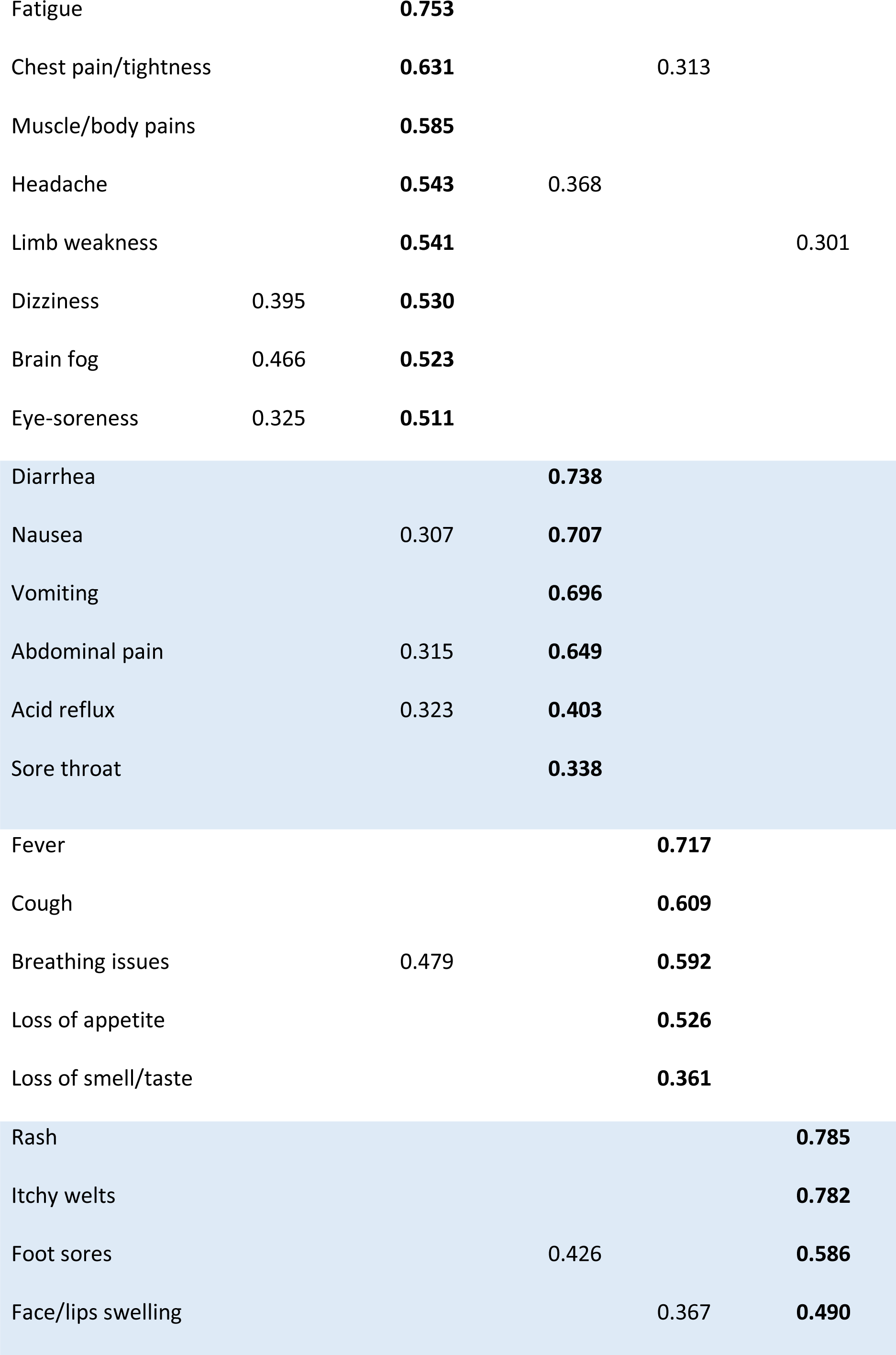
Factors and loadings from the “Initial Symptoms” PCA.

**Table 4.**
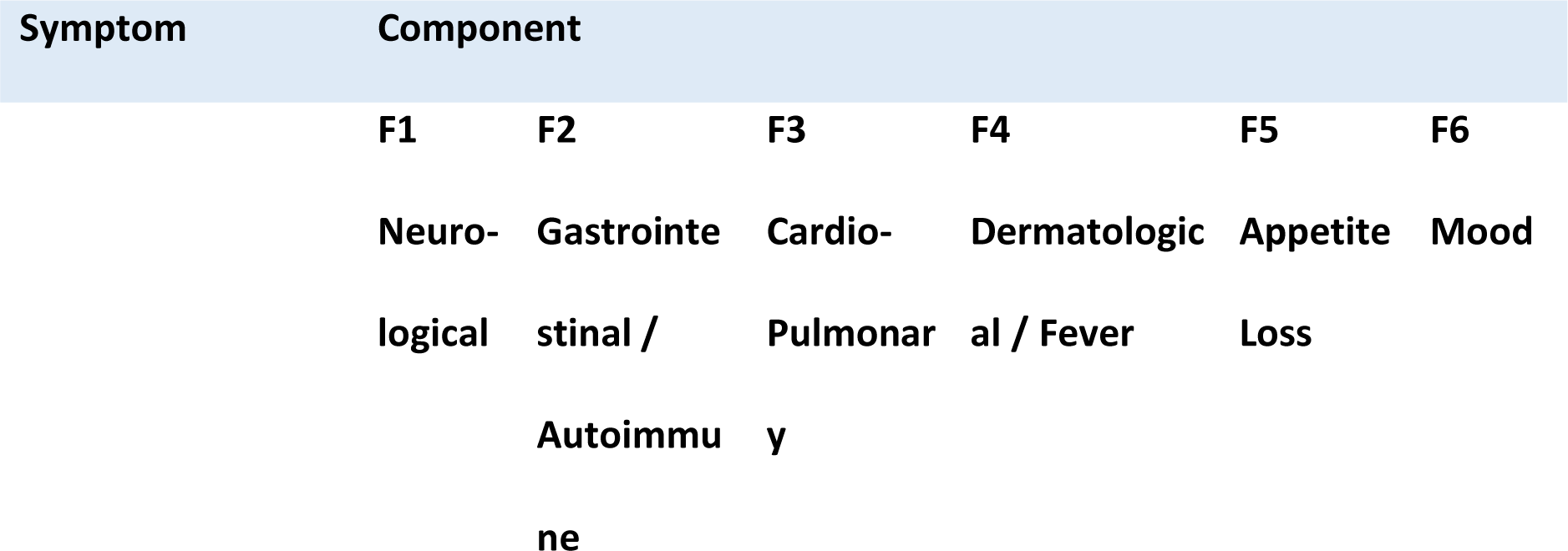
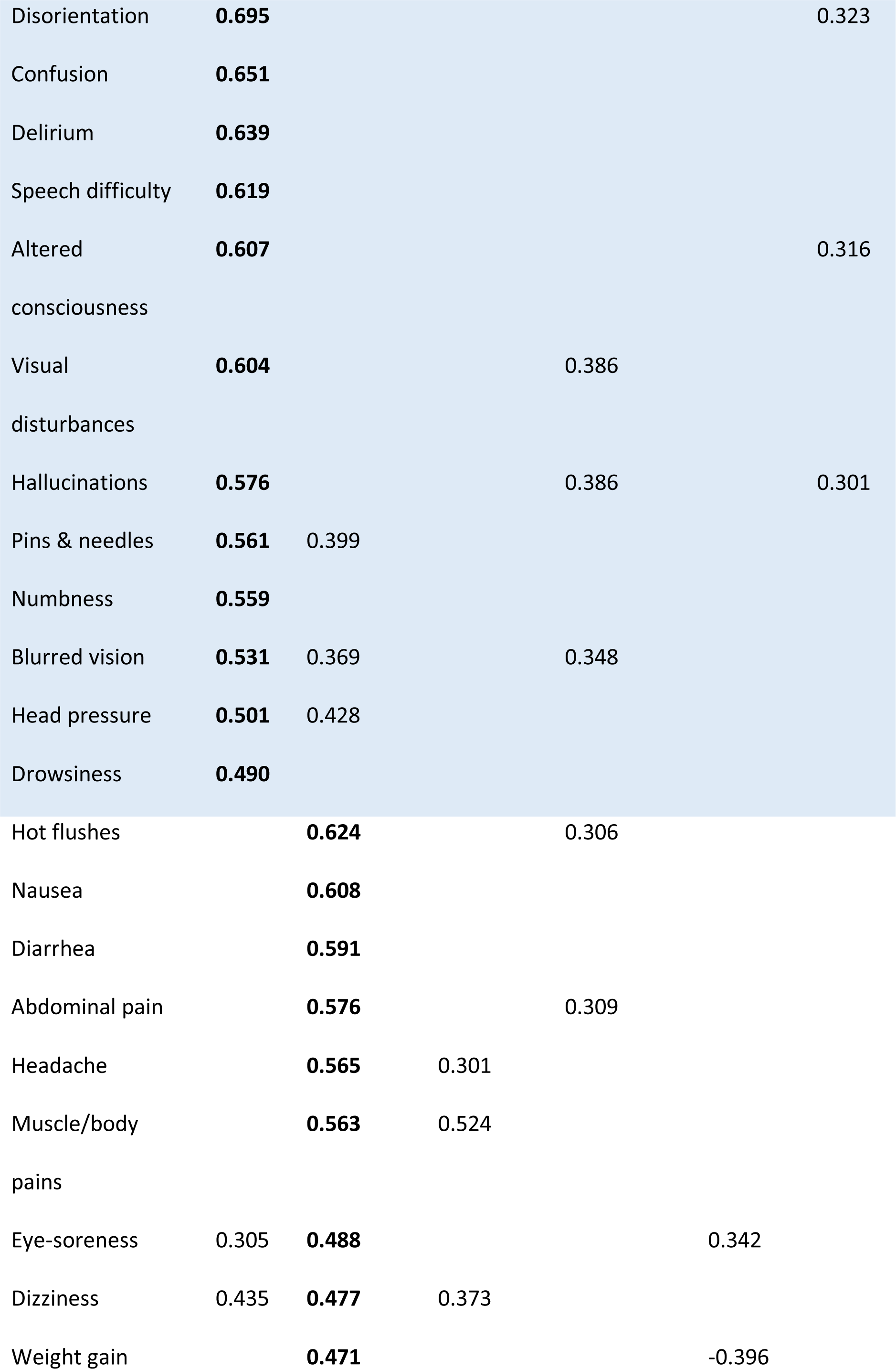
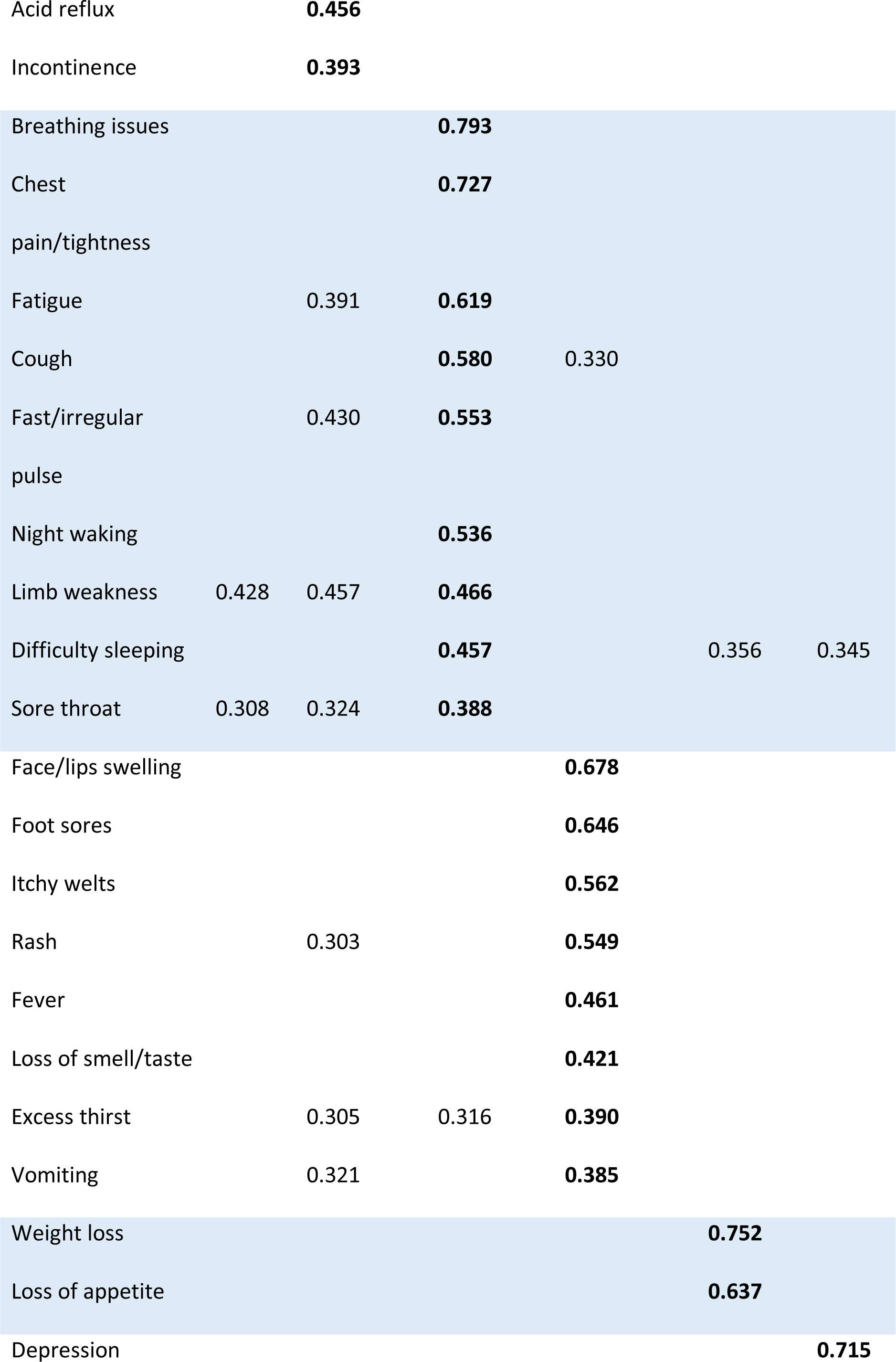
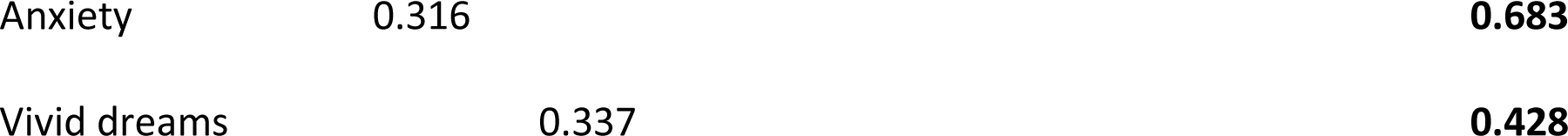
Factors and loadings from the exploratory factor analysis of ongoing “since then” symptoms PCA.

**Table 5.**
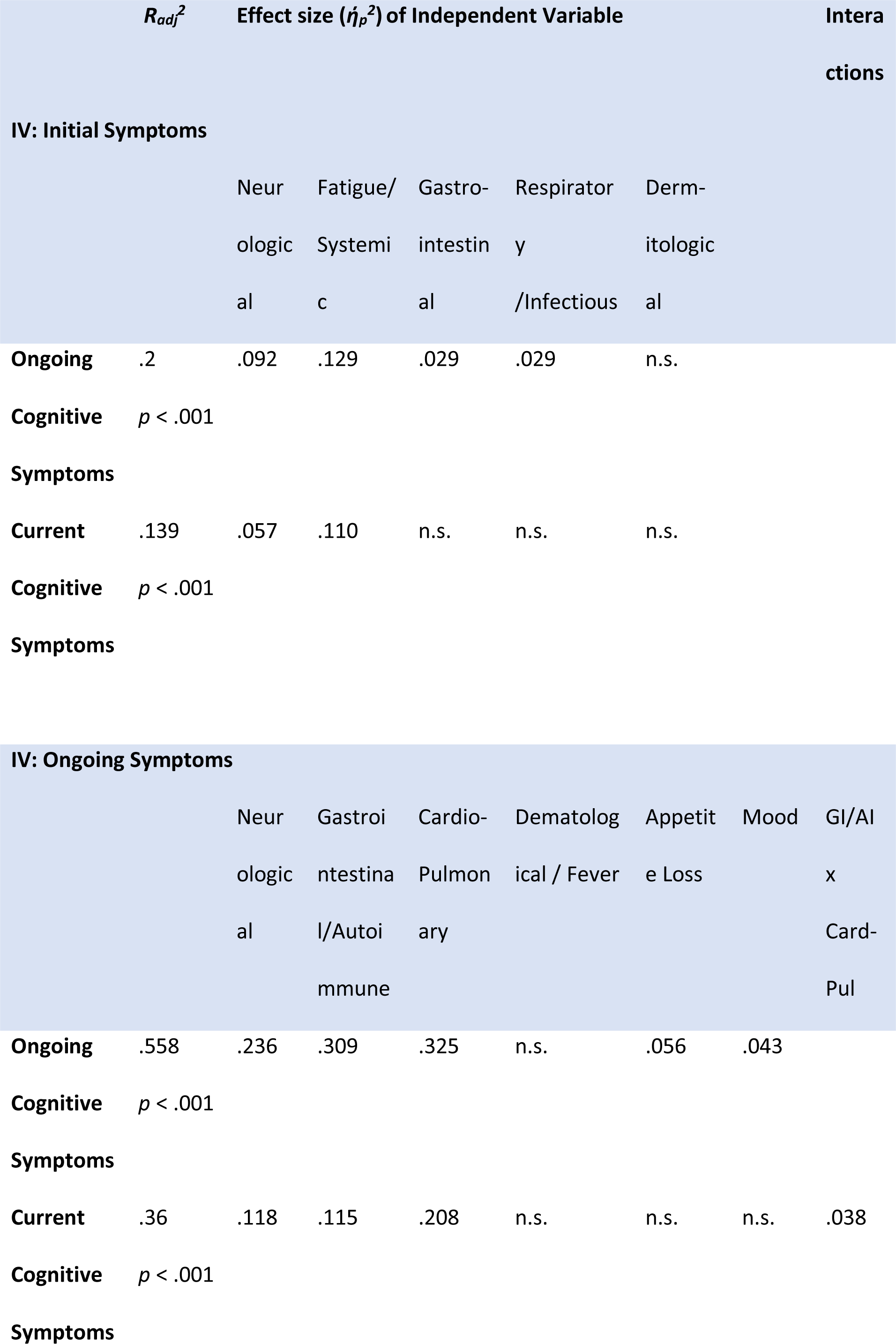
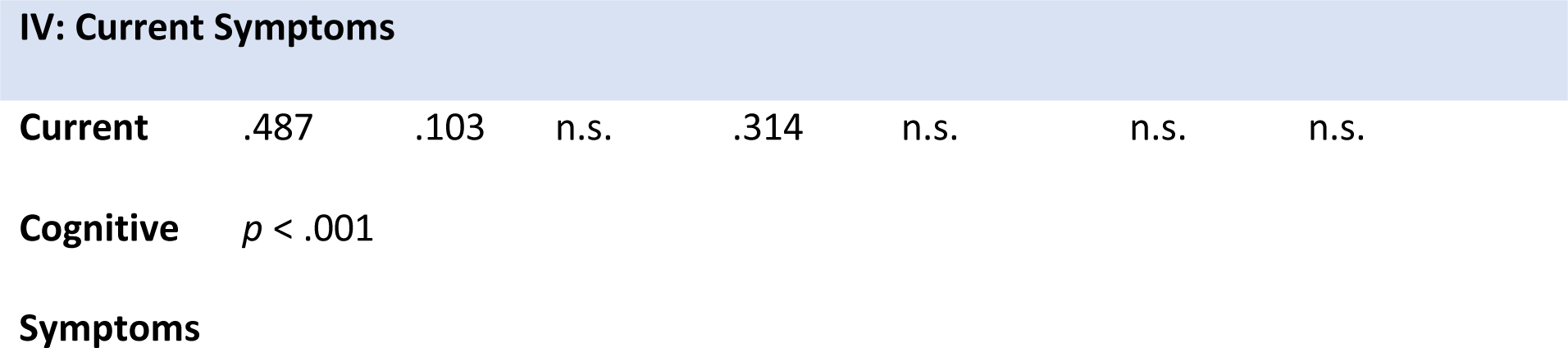
Regression models predicting variation in the cognitive symptom factor (ongoing and current) from non-cognitive symptom factors (initial, ongoing and current). Only partial eta squared (*ήp*) effect size is given here, as beta coefficients are not meaningful for already standardized variables.

**Table 6.**
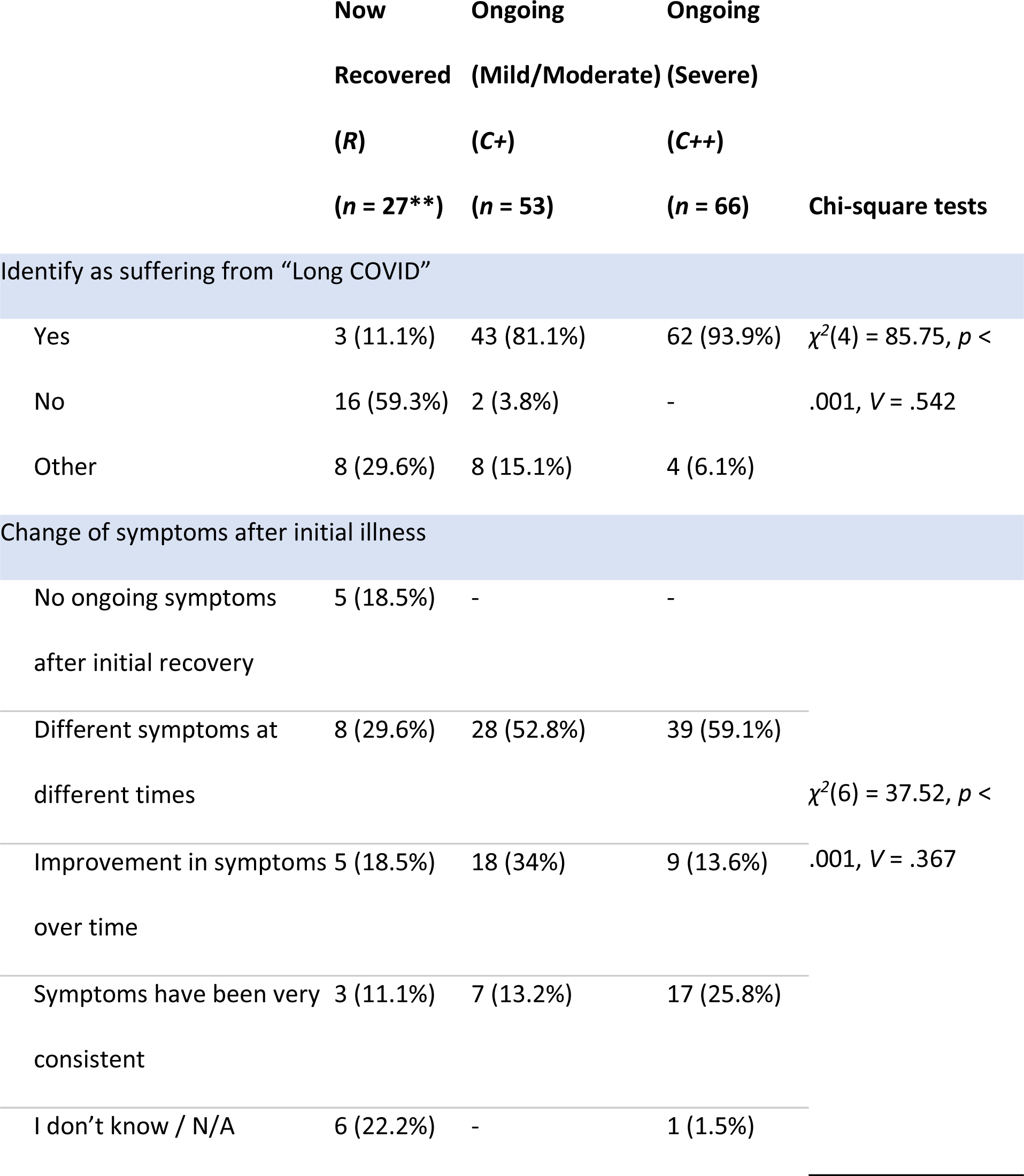
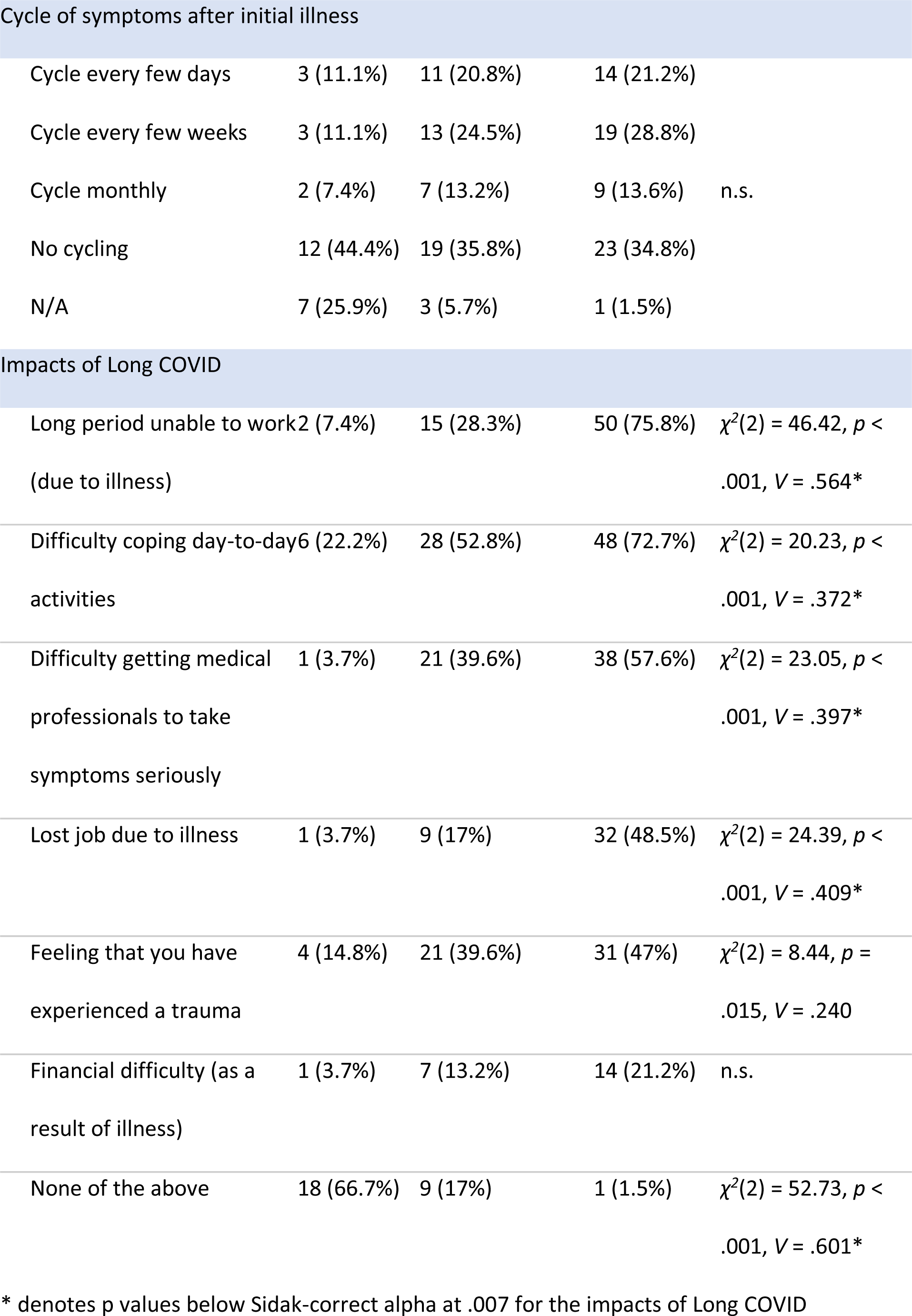
Experiences and impact of Long COVID in different ongoing symptom severity groups.

After controlling for sex, age, education and country, a forward stepwise multinomial logistic regression indicated that no medical history variables predicted COVID-19 progression, however health behaviors including fatty food consumption (*χ^2^*[2] = 23.25, *p* < .001), physical activity (*χ^2^*[2] = 10.31, *p* = .006), and alcohol consumption (*χ^2^*[2] = 8.18, *p* = .017) were all significant predictors. In our sample, people consuming more fatty food had a higher chance of having recovered from COVID-19 (*p* < .001) or having developed mild-moderate ongoing symptoms (*p* < .001) than progressing into severe ongoing symptoms. Higher levels of physical activity were associated with reduced chance of recovery relative to progression onto mild-moderate (*p* = .002) or severe ongoing symptoms (*p* = .034). Those drinking alcohol more frequently were more likely to recover from COVID-19 than to develop severe ongoing symptoms (*p* = .007).

#### 3.2.1 Severity of Initial Illness

The severity of illness in the first 3 weeks of infection was associated with subsequent symptom longevity. Multinomial logistic regression showed that severity of initial illness significantly predicted COVID-19 longevity/severity (*χ^2^*[2] = 24.44, *p* < .001), with higher acute severity associated with more severe subsequent ongoing symptoms (*p*s < .001–.02). This effect was maintained after controlling for sex, age, education and country (*χ^2^*[2] = 12.28, *p* = .002; *C++* > *C+*: *p* = .048; *C++* > *R*: *p* = .001). Those with severe ongoing symptoms experienced more severe initial illness than those whose ongoing symptoms were mild-moderate (*U* = 1258, *p* = .005, Figure 2d) and those who were fully recovered (*U* = 658.5, *p* < .001). The severity difference between the *C+* (Mild/Moderate) subgroup and the *R* subgroup was also significant (*U* = 842, *p* = .034).

Supplementary Table 4 shows the relative frequencies of particular diagnoses received during the acute illness. Of the 109 participants who sought medical assistance, the most common diagnoses received were hypoxia (14.7%), blood clots (5.5%) and inflammation (4.6%).

#### 3.2.2 Symptoms During Initial Illness

Participants were asked to rate a number of specific symptoms they might have experienced during their initial illness from “Not at all” to “Very Severe”. Symptoms that appeared in less than 10% of participants were excluded. Kruskal-Wallis H tests (Sidak *α* = .001) showed significant duration-group differences in 11/33 symptoms in terms of the severity experienced (see Figure 3, more information in Supplementary Table 5). In post-hoc analysis (Sidak *α* = .017), muscle/body pains, breathing issues and limb weakness showed gradation, with the *C++* (Severe) subgroup having experienced the most severe symptoms, followed by the *C+* (Mild/Moderate) subgroup, and the *R* subgroup experiencing the least (*p* ranges < .001–.012). Some symptoms did not show gradation with severity of ongoing symptoms, but were reliably higher in those with ongoing symptoms. Both the ongoing symptoms subgroups reported more severe symptoms of fatigue, brain fog and chest pain/tightness during the initial illness than those that recovered (*p*s </= .001) but did not differ from one another. Those with severe ongoing symptoms experienced more severe nausea and blurred vision than those with mild/moderate or who recovered (*p* ranges < .001–.009). Finally, the *C++* (Severe) subgroup experienced more abdominal pain, altered consciousness and confusion during the initial illness than the *R* subgroup (*p*s </= .001). 

**Figure 3.**
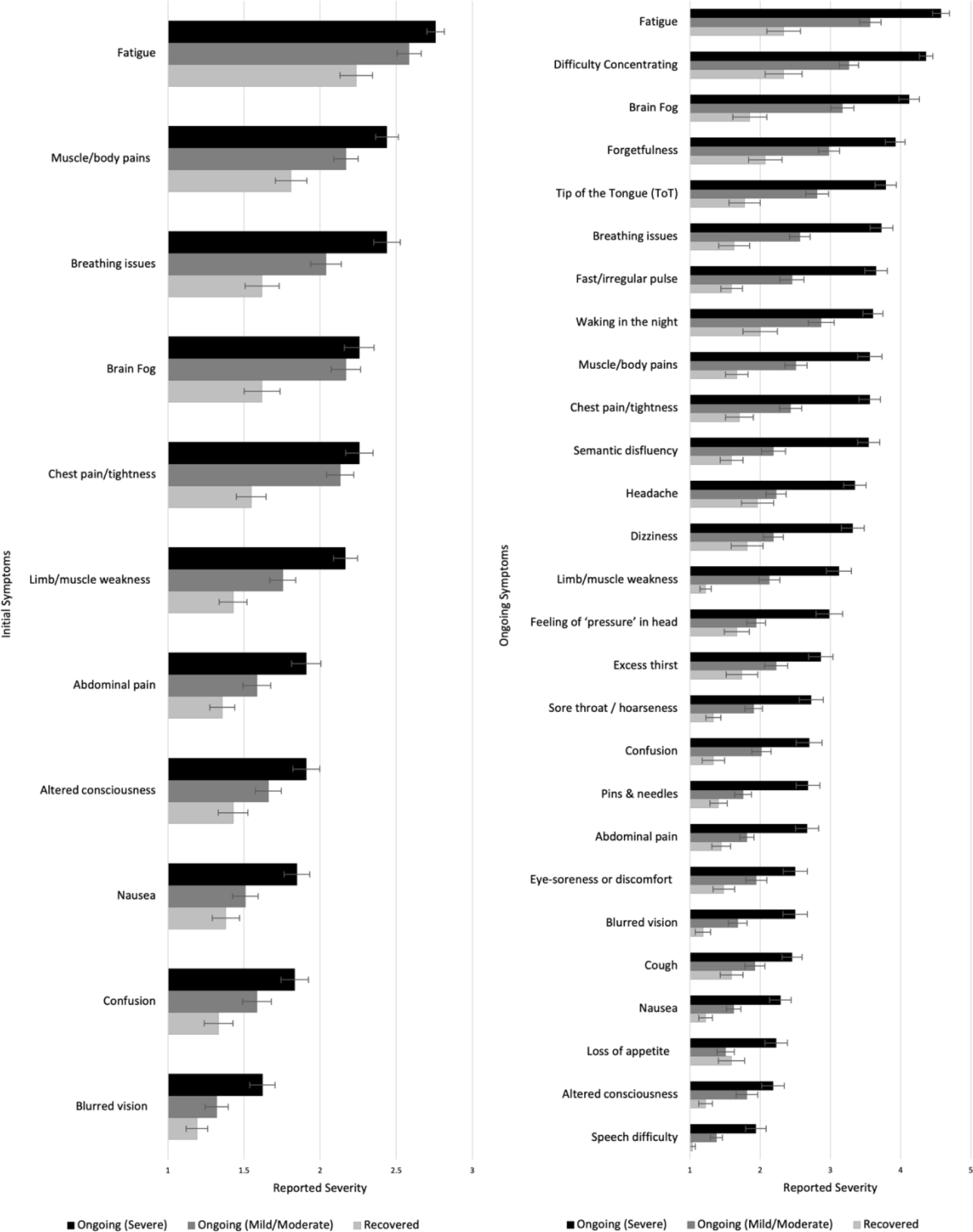
Severity of different symptoms during the initial (left) and ongoing (right) illness among those who recovered or had ongoing mild or severe illness. Higher scores indicate higher severity.

After controlling for sex, age, education and country, a forward stepwise multinomial logistic regression suggested that six initial symptoms significantly predicted COVID-19 progression. These were: limb weakness (*χ^2^*[2] = 25.92, *p* < .001), brain fog (*χ^2^*[2] = 13.82, *p* = .001), chest pain or tightness (*χ^2^*[2] = 10.81, *p* = .005), dizziness (*χ^2^*[2] = 7.82, *p* = .02), cough (*χ^2^*[2] = 7.74, *p* = .021), and breathing difficulties (*χ^2^*[2] = 6.98, *p* = .031). People initially suffering from more severe limb weakness were more likely to experience severe ongoing symptoms than to recover (*p* < .001) or develop mild/moderate ongoing symptoms (*p* < .001). More severe initial breathing issues (*p* = .014) and dizziness (*p* = 0.37) predicted greater likelihood of severe than mild-moderate ongoing symptoms, but people with more severe initial dizziness (*p* = .02) and cough (*p* = .009) were more likely to recover rather than to develop mild/moderate ongoing symptoms. More severe initial brain fog and chest pain/tightness predicted more progression into mild-moderate than either severe ongoing symptoms (brain fog: *p* = .029; chest pain: *p* = .026) or recovery (brain fog: *p* = .001; chest pain: *p* = .007).

#### 3.2.3 Symptoms During Ongoing Illness

Excluding those who reported being totally asymptomatic throughout or feeling completely better very quickly after initial illness (who did not report on ongoing symptoms, *n* = 15), the COVID subgroups were asked to report on their ongoing experience of a list of 52 symptoms (coded here as 1 = *Not at all*, 5 = *Very severe and often*). Symptoms that appeared in less than 10% of participants were excluded. The duration-groups differed significantly in 27/47 symptoms (Sidak *α* = .001; see Figure 3, and Supplementary Table 10). Post-hoc tests (Sidak *α* = .017) showed that the *C++* (Severe) subgroup reported higher levels of severity than the *R* subgroup in all 27 symptoms (*p*s < .001–.017) and than the *C+* (Mild/Moderate) subgroup in all except two (altered consciousness and eye-soreness; *p*s < .001–.017). The *C+* (Mild/Moderate) subgroup also reported experiencing higher severity in 16 symptoms (including fatigue, difficulty concentrating, brain fog and forgetfulness) than the *R* subgroup (*p*s < .001–.016; See Figure 3 and Supplementary Table 10; See also Supplementary Table 11 for similar analysis of current symptoms).

### 3.3 Symptoms in Those with Confirmed or Suspected COVID-19 vs “Other” Illnesses

As much of our sample experienced infection early in the pandemic before widespread testing was available, not all cases included in our COVID group were confirmed by a polymerase chain reaction (PCR) test. Meanwhile, a significant minority of participants had an illness during the pandemic period that they did *not* think was COVID-19 (see Figure 1). We compared symptom prevalence across these three groups (infection status: Unknown, *n* = 55; infection status: Unconfirmed, *n* = 96; infection status: Confirmed, *n* = 65) for both the initial 3 weeks of illness, and the time since then. Those who were still at their first 3 weeks of COVID-19 infection (*n* = 17) and who reported “it is too soon” to comment on their ongoing symptoms (*n* = 3) were not included in this analysis.

The groups significantly differed in 14 out of 31 symptoms during the initial illness (Sidak *α* = .0016; Supplementary Table 6). Both Confirmed and Unconfirmed groups reported higher severity than the Unknown group on 13 symptoms (including fatigue, muscle/body pains and loss of smell/taste; *p* ranges < .001–.014; Sidak *α* = .017). Additionally, the Unconfirmed group reported more severe blurred vision than the Unknown group (*p* < .001), and the Unknown group reported more severe sore throat/hoarseness than the Confirmed group (*p* < .001). As for the differences within those with COVID-19, the Confirmed group experienced greater loss of smell/taste than the Unconfirmed group (*p* = .002), while the Unconfirmed group reported higher levels of breathing issues, chest pain/tightness, sore throat/hoarseness, and blurred vision than the Confirmed group (*p*s = .004–.015).

Of these participants, 177 (Unknown group: *n* = 31; Unconfirmed group: *n* = 88; Confirmed group: *n* = 58) reported experiencing ongoing symptoms after the 3 weeks of illness. Significant group differences were found in 11/47 ongoing symptoms (Sidak *α* = 001; see Figure 4 and Supplementary Table 7). Post-hoc tests (Sidak *α* = .017) showed that, compared with the Unknown group, both the Confirmed and Unconfirmed groups reported higher levels of fatigue, difficulty concentrating, brain fog, tip-of-the-tongue (ToT) problems, muscle/body pains, fast/irregular pulse, semantic disfluency, chest pain/tightness, limb weakness and loss of smell/taste (*p*s </= .001). The Unconfirmed group also experienced higher level of night waking (*p* = .001) than the Unknown group. There were no significant differences in ongoing symptoms between the Confirmed and the Unconfirmed groups.

**Figure 4.**
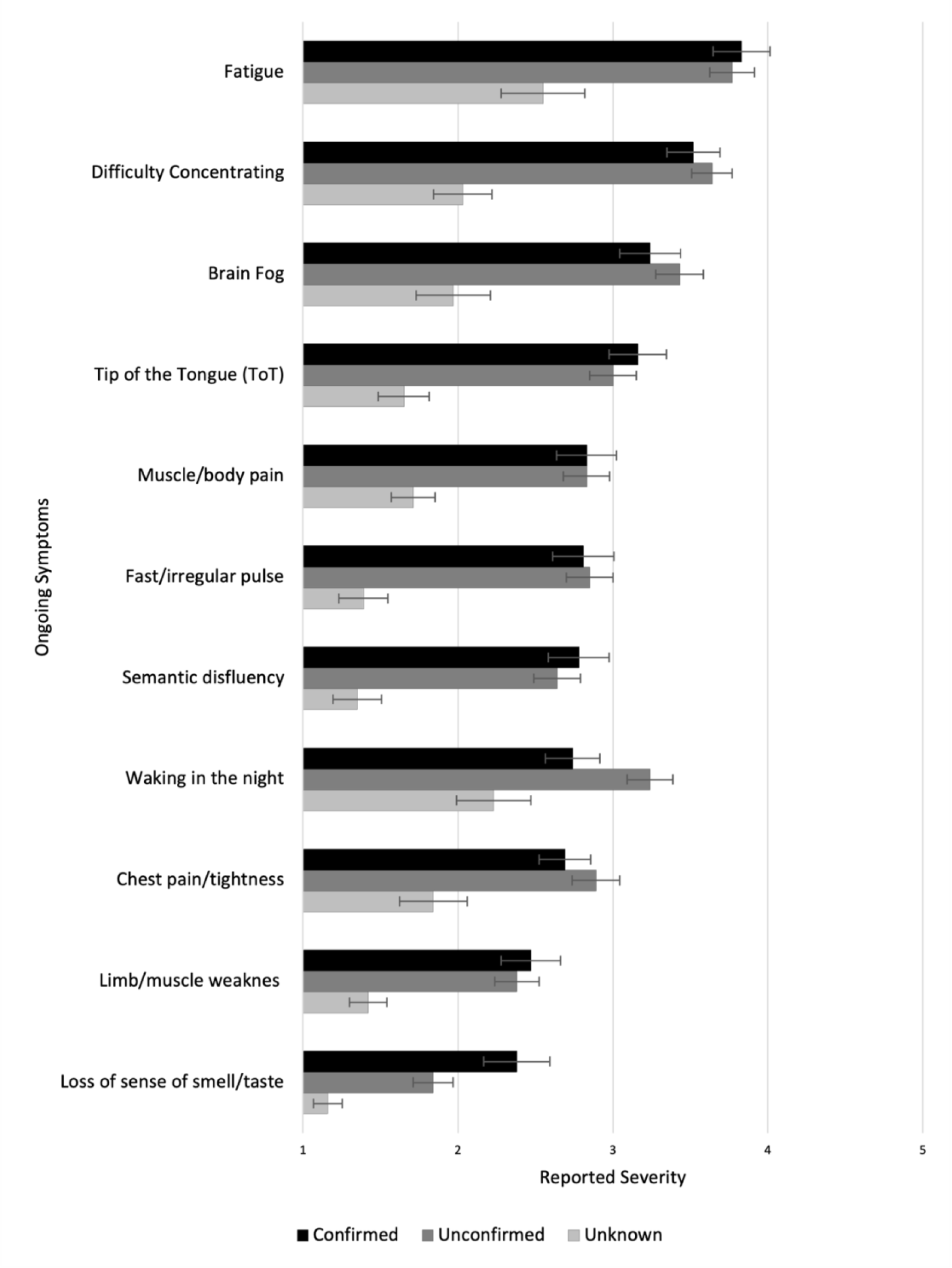
Experience of ongoing symptoms in Unknown, Unconfirmed COVID, and Confirmed COVID groups.

### 3.4 Characterizing Symptom Profiles

While data on individual symptoms are useful in identifying highly specific predictors, these are too numerous for more systematic analysis, which require data-reduction. A stated aim of this study was to identify symptom profiles that may be informative as to underlying pathology. To determine suitable groups of symptoms, we employed exploratory principal component analysis (PCA) with varimax rotation. Based on our high number of items (Nunnally 1978) and the novelty of the subject (Henson and Roberts 2006), we performed two PCAs, one for the initial symptoms and another one symptoms experience since the initial phase. We then used the high-loading items on the “since then” symptom factors to calculate profiles for currently experienced symptoms.

#### 3.4.1 Initial Symptom Factors

To group the initial symptoms, we included 34 symptoms in the PCA after excluding paralysis and seizures (experienced by less than 10% of the participants). A total of 164 participants reported on their symptoms during the first 3 weeks of illness using a 3-point scale: *Not at all, Yes, mildly, Yes, very severe*. The Kaiser-Meyer-Olkin (KMO) test (value 0.861) and Bartlett’s test of sphericity (*χ^2^*[528] = 2250, *p* < .001) showed the data were suitable for factor analysis. We employed the varimax rotation. Initially, nine factors were obtained with eigenvalue > 1.0, which was reduced to five via Cattell’s Scree test (Kline 2013). Assessments were conducted of 4, 5 and 6 factor solutions for interpretability and robustness. The ratio of rotated eigenvalue to unrotated eigenvalue was higher for the 5-factor solution than for the 4- or 6-factor solutions, and this structure was also the most interpretable. We thus proceeded with a 5-factor solution, which explained 50.59% of item variance with last rotated eigenvalue of 1.998.

We labelled the new components as “F1: Neurological”, “F2: Fatigue/Systemic”, “F3: Gastrointestinal”, “F4: Respiratory/Infectious”, and “F5: Dermatological”. We computed the factor scores using the regression method.

People who went on to experience ongoing symptoms showed higher factor scores in the Fatigue/Systemic symptom factor during the initial illness (*F*[2,158] = 23.577, *p* < .001), but did not differ in any other initial symptom factor. Pairwise analysis revealed that those who recovered were significantly less likely to experience Fatigue/Systemic symptoms than those with mild-moderate (*p* < .001) or severe (*p* < .001) ongoing symptoms (Figure 5).

**Figure 5.**
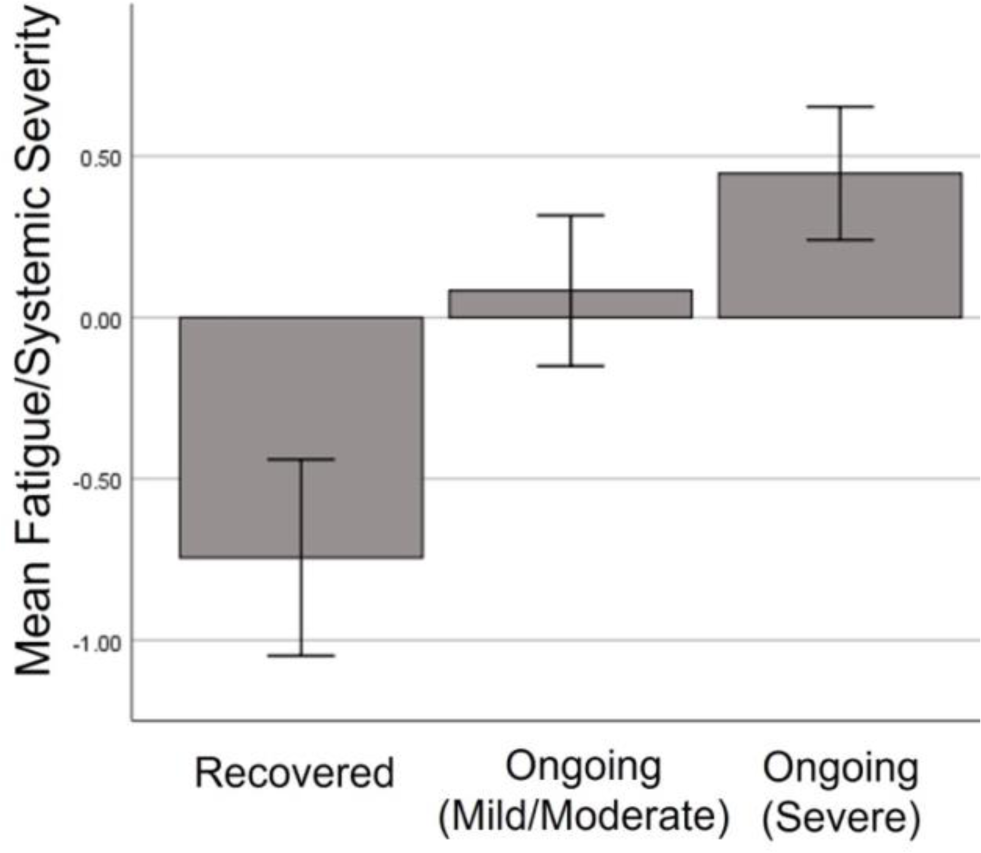
Severity of Fatigue/Systemic symptom factor during initial illness among those who went on to full recover, or have ongoing mild or severe symptoms.

#### 3.4.2 Ongoing Symptom Factors

We performed a second PCA (*N* = 149) using the symptoms experienced since the acute phase (after the first 3 weeks), including 47 symptoms. Paralysis, seizures, face/lips swelling, hallucinations, and delirium were excluded (experienced by less than 10% of the participants). A total of 149 participants reported on their symptoms over the time since the first 3 weeks of illness using a 5-point scale: 1 = *Yes, very severe and often*, 2 = *Yes, very severe and occasional*, 3 = *Yes, mildly and often*, 4 = *Yes, mildly and occasional*, and 5 = *Not at all*. The KMO test (value 0.871) and Bartlett’s test of sphericity (*χ^2^*[861] = 3302, *p* < .001) showed suitability for factor analysis. We employed the varimax rotation. PCA showed 11 components with eigenvalues > 1.0, and this was reduced to 6 via inspection of the eigenvalue gradient (scree plot). The ratio of rotated eigenvalue to unrotated eigenvalue was higher for the 7-factor solution, followed by the 6-factor. The 6- and 7-factor solutions were differentiated by subdivision of the second factor, reducing the degree of cross-loading. However, the 7-factor solution was less interpretable and less robust to removal to cross-loaders (the presence of which can be accepted from a pathology perspective, given that multiple mechanisms can produce the same symptom). As such, we proceeded with the 6-factor solution, which explained 54.17% of item variance and had a last rotated eigenvalue of 2.227.

We labelled the new components as “F1: Neurological”, “F2: Gastrointestinal/ Autoimmune”, “F3: Cardio-Pulmonary”, “F4: Dermatological/Fever”, “F5: Appetite Loss” and “F6: Mood”. We computed the factors’ scores using the regression method.

In order for cognitive symptoms (brain fog, forgetfulness, tip-of-the-tongue (ToT) problems, semantic disfluency and difficulty concentrating) to be used as a dependent variable, these were isolated and a PCA run separately. A single component emerged, with all the cognitive symptoms loading homogeneously highly (see Supplementary Table 8). The KMO test (value 0.886) and Bartlett’s test of sphericity (*χ^2^*[10] = 564, *p* < .001) indicated suitability for factor analysis, and the single 5-item factor explained 76.86% of variance.

#### 3.4.3 Current Symptoms

The current symptoms assessed were the same as the ongoing symptoms, but rated dichotomously as either currently present or absent. To estimate the degree to which current symptoms aligned with the factors established for the ongoing period, we generated a quasi-continuously distributed variable according to how many of the high loading (>/= 0.5) items from the ongoing factors were recorded as present currently. Using this *sum scores by factor* method (Hair 2009; Tabachnick, Fidell, and Ullman 2007), each score was subsequently divided by the number of items in that factor producing quasi “factor scores” that were comparable and indicative of “degree of alignment” of current symptoms to established factors.

To assess the stability and specificity of symptom profiles between these periods, serial correlations were conducted for corresponding and non-corresponding factors. Correlations of the same factor across time points were materially higher (> 0.2) from the next highest correlation among the 5 non-corresponding factors, with Williams tests (Steiger 1980) giving the narrowest gap at *p* = .003 (Neurological: *r* = .676, *t* = 5.712; Gastrointestinal/Autoimmune: *r* = .531, *t* = 3.778; Cardio-Pulmonary: *r* = .678, *t* = 7.272; Dermatological/Fever: *r* = .523, *t* = 3.364; Appetite Loss: *r* = .591, *t* = 5.017; Mood: *r* = .490, *t* = 4.803). This consistency suggests that while particular symptoms may fluctuate, the profile of symptoms—once grouped into an adequately supported factor—is moderately stable for individuals, and can be relatively well represented by a “snapshot” of current symptoms. For completeness, an additional factor analysis was conducted on the current symptoms, which are reported in Supplementary Table 12.

One symptom factor showed change over time since infection, suggesting higher severity in those who had been ill for longer: Number of weeks since infection (positive test / first symptoms) was positive correlated with severity of ongoing severity of Cardio-Pulmonary symptoms (*r*[147] = .271, *p* < .001; Figure 6) and, to a weaker extent, current alignment with the same factor (*r*[147] = .206, *p* = .012) however only the former association survived correction for multiple comparisons (Sidak *α* = .0085).

**Figure 6.**
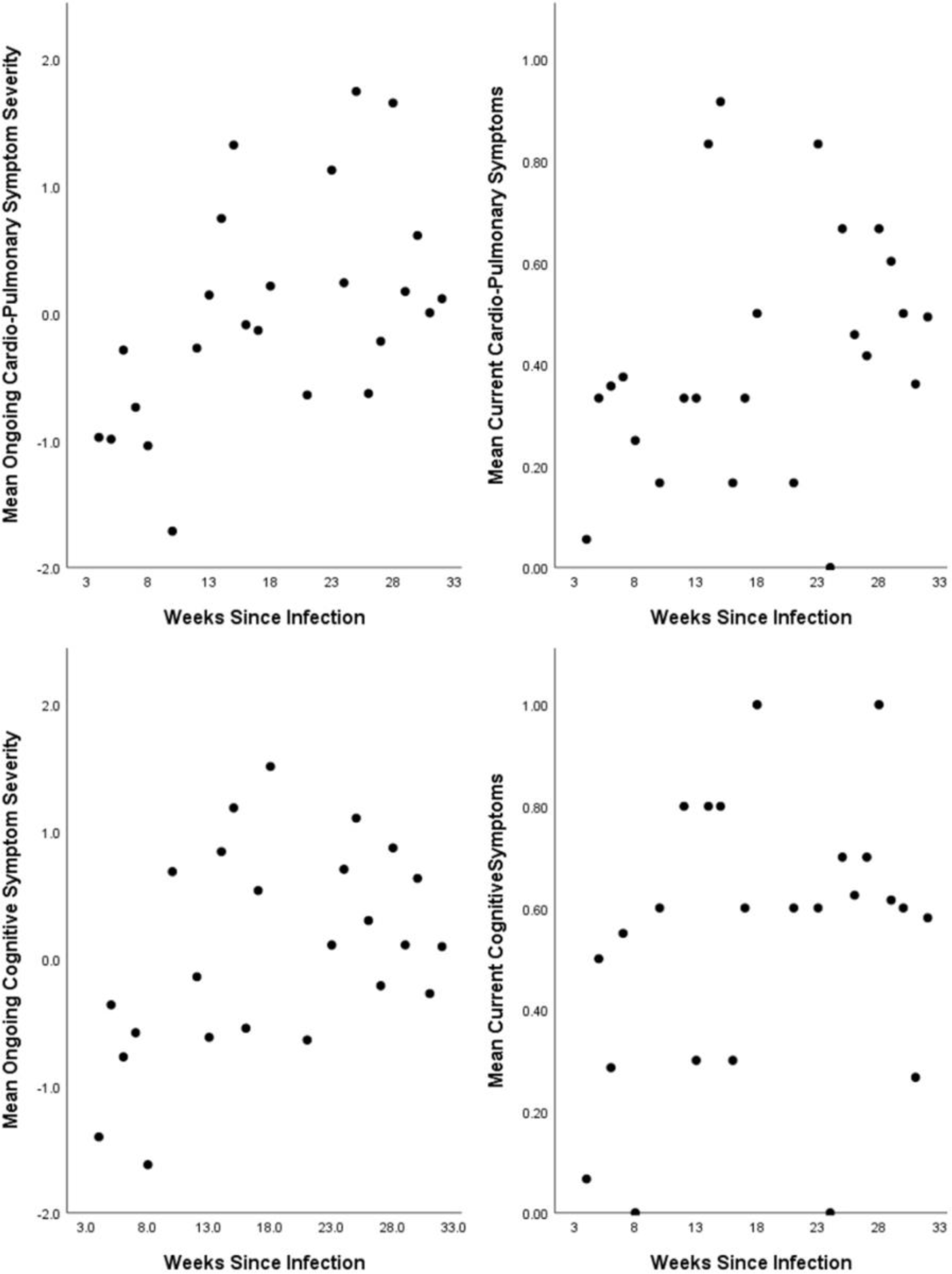
Association between number of weeks since infection and severity of (top) Cardio-Pulmonary Symptoms and (bottom) cognitive symptoms in the entire period since the initial infection (left) and the past 1–2 days (right). Higher scores indicate higher symptom severity.

#### 3.4.4 Cognitive Symptoms

Within those currently experiencing symptoms (*n* = 126), 77.8% reported difficulty concentrating, 69% reported brain fog, 67.5% reported forgetfulness, 59.5% reported tip-of-the-tongue (ToT) word finding problems and 43.7% reported semantic disfluency (saying or typing the wrong word).

Symptoms experienced during the initial illness significantly predicted both ongoing and current cognitive symptoms (Figure 7). A linear regression with backwards deletion found that the best model contained the Neurological, Fatigue/Systemic, Gastrointestinal and Respiratory/Infectious symptom factors and predicted 20% of variance (*p* < .001). Table 5 shows that the Fatigue/Systemic symptoms factor (*ή_p_^2^* = .129) was the better predictor followed by the Neurological symptom factor (*ή_p_^2^* = .092). For current cognitive symptoms, the best model contained both the Neurological and Fatigue/Systemic symptom factors, together predicting 13.9% of variance. (*p* < .001). Of the two, the Fatigue/Systemic factor was the better predictor (*ή_p_^2^* = .110). No interactions between factors contributed significantly and were thus not included in the final models.

**Figure 7.**
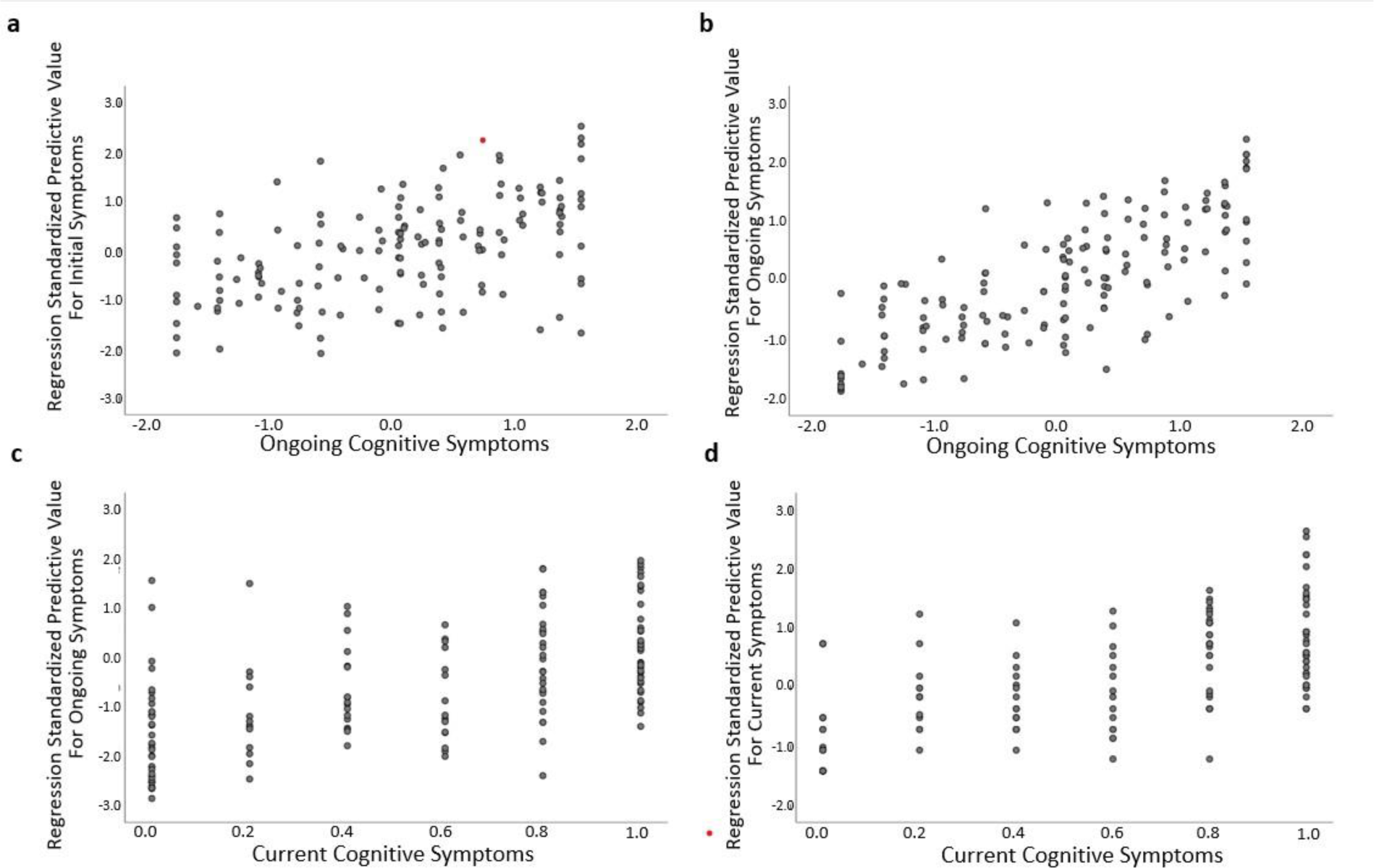
Association between combined regression model predicted value for a) initial symptom factors and ongoing cognitive symptoms; b) initial symptom factors and current cognitive symptoms; c) ongoing symptom factors and ongoing cognitive symptoms and d) current symptom factors and current cognitive symptoms.

A similar, but much stronger, pattern emerged when considering the predictive value of ongoing (non-cognitive) symptoms (see Figure 7). Using backwards elimination to factors with significance (*p* < .05), all factors except dermatological/fever remained in the model, which predicted over 55% of variance (*R_adj_^2^* = .558, *p* < .001). The effect size (*ή_p_^2^*) for each factor is given in Table 5. The Gastrointestinal/Autoimmune and Cardio-Pulmonary factors were the biggest contributors to the model. Indeed, in an extreme elimination model in which contributing factors were limited to two or fewer, these two factors alone predicted 38% of variance retaining strong significance (*p* < .001). No interactions between factors contributed significantly and were thus not included in the final models. Ongoing symptoms also predicted current cognitive symptoms. The best model predicted 36% of the variance (*p* < .001) and included the Neurological, Gastrointestinal/Autoimmune and Cardio-Pulmonary factors and an interaction between the Gastrointestinal/Autoimmune and Cardio-Pulmonary factors. Of these, Cardio-Pulmonary symptoms were the strongest predictor (*ή_p_^2^* = .208), with neurological (*ή_p_^2^* = .118) and Gastrointestinal/Autoimmune (*ή_p_^2^* = .115) being relatively equal.

Current symptom factors also strongly predicted current cognitive symptoms. The backwards elimination model left two contributing factors: Neurological and Cardio-Pulmonary. Together these predicted around 50% of variance (*R_adj_^2^* = .487). Of these, Cardio-Pulmonary was the stronger predictor (*ή_p_^2^* = .314). Indeed, when the model was limited to just this factor, this model still predicted 43% of the variance.

There was a significant association between degree of cognitive symptoms and duration of illness. Those who had been ill for longer were more likely to report having had cognitive symptoms throughout the ongoing illness (*r*[147] = .262, *p* = .001) and to be experiencing them at the time of test (*r*[147] = .179, *p* = .03) (Figure 6).

### 3.5 Experiences and Impact of Long COVID

Here we limited analysis to all those who reported some degree or period of ongoing symptoms following COVID-19 (i.e., excluding those who reported being totally asymptomatic throughout or feeling completely better very quickly after initial illness (*n* = 15). Of the remaining 146 participants, 108 (74%) self-identified as suffering or having suffered from “Long COVID”.

We examined the impact and experiences of ongoing illness. In most cases, the nature and degree of negative experience of ongoing symptoms scaled with perceived severity. The change in symptoms over time differed between severity subgroups (*χ^2^*[6] = 37.52, *p* < .001, *V* = .367). The *C++* (Severe) subgroup were more likely to report that symptoms were consistent over time, while those with mild-moderate ongoing symptoms were more likely to report improvement in symptoms. As might be expected, the *R* subgroup were alone in reporting complete resolution of symptoms after recovery from the initial illness (Supplementary Table 9).

Long COVID has significant impact on individuals’ lives. Over 54.6% of those with ongoing symptoms had experienced long periods unable to work and 34.5% had lost their job due to illness, 63.9% reported difficulty coping with day-to-day activities, 49.6% had had difficulty getting medical professionals to take their symptoms seriously, and 43.7% felt that they had experienced a trauma, while 17.6% had experienced financial difficulty as a result of illness. These impacts scaled with symptom severity. Those with severe ongoing symptoms were more likely to report being unable to work for a long period due to illness (*χ^2^*[2] = 46.42, *p* < .001, *V* = .564), having difficulty coping with day-to-day requirements (*χ^2^*[2] = 20.23, *p* < .001, *V* = .372), having difficulty getting medical professionals to take their symptoms seriously (*χ^2^*[2] = 23.05, *p* < .001, *V* = .397), and losing their job due to illness (*χ^2^*[2] = 24.39, *p* < .001, *V* = .409). In contrast, the *R* subgroup tended to report experiencing none of the above (*χ^2^*[2] = 52.73, *p* < .001, *V* = .601).

We further compared job-loss with the No COVID group (*n* = 185). Those with ongoing symptoms were more likely to have lost their job than those who had not experienced COVID-19 (*χ^2^*[1] = 26.74, *p* < .001, *V* = .297). The most common reason for job-loss among those with ongoing symptoms was illness (*χ^2^*[1] = 56.85, *p* < .001, *V* = .432), while the most common reason in the No COVID group was economy (*χ^2^*[1] = 7.67, *p* = .006, *V* = .159).

## 4 Discussion

Here we report the initial findings from a cross-sectional/longitudinal study investigating cognition post-COVID-19.

One aim of this first publication was to characterize the “COVID and Cognition Study” sample. Within the COVID group, we recruited specifically to get good representation of those who were experiencing or had experienced ongoing symptoms. Indeed, 74% identified with the term “Long COVID”. Our final sample had a relatively even spread of those that had fully recovered at the time of test (42), or had mild-moderate (53) or severe (66) ongoing symptoms. Medical history did not differ between those experiencing ongoing symptoms and those who recovered, and indeed in terms of health behaviors those with ongoing symptoms were in general “healthier”, being more likely to have previously been regular consumers of fatty or sugary food, and more likely to have been physically active and consumers of fruit and vegetables.

The nature of the initial illness was found to have a significant impact on the likelihood and severity of ongoing symptoms. Despite this sample almost entirely comprised of non-hospitalized patients, those with more severe initial illness were more likely to have ongoing symptoms, and for those symptoms to be more severe. This suggests even in “community” cases, acute infection severity is a predictor of vulnerability to Long COVID. In an analysis of all symptoms experienced during the initial acute illness, there were several that were predictive of presence or severity of ongoing symptoms. In particular, individuals with severe ongoing symptoms were significantly more likely to have experienced limb weakness during the initial illness than those that recovered.

We asked participants to retrospectively report on a large number of symptoms over three time periods: acute illness (first 3 weeks), ongoing illness (“since then”—i.e., the time since the initial infection), and currently experienced (past 1–2 days). Given the highly heterogenous nature of Long COVID, we used principal component analysis (PCA) with the aim to ascertain whether there may be different phenotypes of the condition within our sample—that is to say, that there may be certain types of symptoms that tend to (or not to) co-occur.

We identified 5 factors for the initial illness. These included a “Neurological” factor which was characterized by disorientation, delirium and visual disturbances, a “Fatigue/Systemic” factor which was characterized by fatigue, chest pain/tightness and muscle/body pains, a “Gastrointestinal” factor, characterized by diarrhea, nausea and vomiting, a “Respiratory/Infectious” factor characterized by fever, cough, and breathing issues and a “Dermatological” factor characterized by rash, itchy welts, and foot sores. Those who went on to have ongoing symptoms were more likely to have symptoms aligned with the “Fatigue/Systemic” factor.

For symptoms experienced during the ongoing illness, 6 factors were identified. The “Neurological” factor was characterized by disorientation, confusion and delirium. The “Gastrointestinal/Autoimmune” factor was characterized by hot flushes, nausea and diarrhea.

The “Cardio-Pulmonary” factor was characterized by breathing issues, chest pain/tightness and fatigue. The “Dermatological/Fever” factor was characterized by face/lips swelling, foot sores, and itchy welts. The “Appetite Loss” factor was characterized by weight loss and loss of appetite. Finally, the “Mood” factor was characterized by depression, anxiety and vivid dreams.

For both the acute and ongoing illness, these symptom factors resemble those found in previous studies (e.g., Ziauddeen et al. 2021), with some quite coherent cardio-pulmonary clusters, and other less specific “multi-system” profiles which may reflect more systemic issues such as inflammation, circulation, or endocrine function. One international web-based survey on 3762 respondents traced 66 symptoms and categorized them into 10 organ systems. The researchers then grouped symptoms according to their time-course and found that affecting the same organ system change differently over time, suggesting that there is an evolution of symptomatology as illness progresses (Davis et al. 2021). Finally, the REACT study (Whitaker et al. 2021) extracted two stable clusters at 12 weeks, incorporating a “tiredness cluster” (characterized by tiredness, muscle aches, difficulty sleeping and shortness of breath) and a “respiratory cluster” (characterized by shortness of breath, tight chest and chest pain).

### 4.1 Predictors of Cognitive Difficulties

A large proportion of our sample reported cognitive difficulties. We isolated the cognitive symptoms for the ongoing and current illness and computed a single factor including only these. Although this cognitive variable contained only 5 items, in their quantified form these gave a continuous distribution with minimal skew allowing us to use this as a dependent variable. Using this, we investigated which (non-cognitive) symptom factors during both the initial and ongoing illness predicted significant variance in severity of cognitive symptoms.

Fatigue/Systemic, Neurological, Gastrointestinal and Respiratory/Infectious symptoms during the initial illness together predicted around 20% of variance in ongoing (“since then”) cognitive symptoms, and a similar model (containing only Neurological and Fatigue/Systemic symptoms) predicted nearly 14% of variance in current cognitive symptoms.

These findings strongly suggest that experience of neurological symptoms during the acute illness are significant predictors of self-reported cognitive impairment. While only one factor is named “Neurological” both this and the Fatigue/Systemic factor contain clear elements of neurological involvement. Indeed, headache, dizziness, and brain fog all loaded more highly on the Fatigue/Systemic factor than on the Neurological factor (which was more characterized by disorientation, visual disturbances, delirium and altered consciousness). This suggests different types of neurological involvement, potentially reflecting neuroinflammation (the Fatigue/Systemic factor) and encephalitis (the Neurological factor) respectively. It is of note then that both these factors independently predicted subjective cognitive problems. It will be an important next step in the investigation to explore whether these two factors differentially predict performance on tests assessing different cognitive abilities.

Participants’ experience of ongoing Neurological, Cardio-pulmonary, Gastrointestinal/Autoimmune, Mood and Appetite Loss symptom factors all predicted current cognitive symptoms, together explaining around over 55% of variance. Unlike the initial symptom factors, the vast majority of neurological symptoms were contained within the Neurological factor for ongoing symptoms, with only headache and dizziness loading more strongly into the Gastrointestinal/Autoimmune factor. This latter factor was instead more characterized by symptoms associated with systemic illness—potentially endocrine, or reflecting thyroid disruption—including diarrhea, hot flushes and body pains. An additional predictor here was Cardio-Pulmonary symptoms, a factor which was quite narrowly characterized by symptoms associated with breathing difficulties. Alone, the Gastrointestinal/Autoimmune and Cardio-Pulmonary factors explained a large proportion of the variance (36%), suggesting these were the biggest contributor to individual differences in cognitive symptoms. These findings suggest that the symptoms linked with cognitive issues are not so specifically neurological as during the initial illness, but may also incorporate problems with heart and lung function (potentially implying hypoxia) and with other ongoing ill health that is harder to label (resembling symptoms of the menopause, Crohn’s disease, hypothyroidism, and a number of other conditions). Again, these associations align with previous findings, in which cardio-pulmonary and cognitive systems clustered in the same factor (Ziauddeen et al. 2021).

In terms of current symptoms, the Cardio-Pulmonary factor again emerged as a significant predictor, this time paired with Neurological symptoms and predicting nearly 50% of variance. It is potentially notable that both the cognitive and Cardio-Pulmonary factors showed positive correlation with length of illness, suggesting either that the same disease process underpinning both increases in severity over time, or that the relationship between the two may be the result of both being symptoms more commonly still experienced in those with longer-lasting illness. Longitudinal investigation within individuals would be necessary to disambiguate this.

### 4.2 Importance

Of those experiencing Long COVID, more than half (and 75% of those with severe symptoms) reported long periods unable to work. Indeed, 35% (∼50% of those with severe ongoing symptoms) had lost their jobs or were unable to work due to illness, with job losses being significantly higher in this group compared to the No COVID group (who had also experienced some unemployment, but due to other factors). These findings chime with evidence from other studies on Long COVID (e.g., Davis et al. 2021; Ziauddeen et al. 2021). Notably, Davis and colleagues (2021) found that in their sample 86% of participants reported that it was the cognitive dysfunction *in particular* that was impacting their work—with 30% reporting this leading to “severe” inability to function at work.

These findings are of particular concern given the prevalence of Long COVID as a percentage of the workforce. The pandemic and associated lockdowns have had an unprecedented impact on national economies and individual livelihoods (Jones, Palumbo, and Brown 2021), where the global economy has experienced the worst decline since the Great Depression (IMF 2020) and global unemployment has increased by over 30 million (ILO 2021).

The reported experiences of those with Long COVID—many of whom were at least 6 months into their illness at the time of completing the study—suggest that in addition to these broad economic challenges, society will face a long “tail” of workforce morbidity. It is thus of great importance both not just for the sake of individuals, but for broader society, to be able to prevent, predict, identify and treat issues associated with Long COVID. However, a major roadblock to this is clinicians not having the information or experience with which to offer assistance. A significant number (over 50% of those with severe symptoms) of our sample reported struggling to get medical professionals to take their symptoms seriously. Part of this issue will be the nature of the symptoms experienced. Patients whose symptoms cannot be, or are not routinely, clinically measured (such as cognitive symptoms; Kaduszkiewicz et al. 2010) are at greater risk of “testimonial injustice”—that is, having their illness dismissed by medical professionals (De Jesus et al. 2021). The novel and heterogenous nature of Long COVID also provides a particular challenge for clinicians dealing with complex and undifferentiated presentations and “medically unexplained symptoms” (Davidson and Menkes 2021). The data presented here demonstrate that cognitive difficulties reported by patients can be predicted by severity and pattern of symptoms during the initial stages of infection, and during the ongoing illness. These findings should provide the foundation for clinicians to assess the risk of long-term (6 months+) cognitive difficulties, as well as for researchers to investigate the underlying mechanism driving these deficits. In our next paper, we will explore the association between general and cognitive symptoms and performance on cognitive tasks, with the aim of establishing whether self-reported cognitive issues translate into “objective” deficits on cognitive evaluations.

Some have argued that cognitive changes following COVID-19 infection may reflect changes related to experience of lockdown. There is indeed some evidence that pandemic-related lifestyle changes impact cognition (e.g., Okely et al. 2021). However, many of these studies did not record COVID-19 infection history (Okely et al. 2021; Smirni et al. 2021) so it is difficult to ascertain to what degree these findings may have been related to COVID-19 infection. One study that did control for this (Fiorenzato et al. 2021) identified significant declines in self-reported attention and executive function, however showed reduced reports of forgetfulness compared with pre-lockdown. Our results show that, compared to individuals who experienced a (probable) non-COVID-19 illness during the pandemic, those with suspected or confirmed COVID-19 infection experienced greater levels of fatigue, difficulty concentrating, brain fog, tip-of-the-tongue (ToT) word finding problems and semantic disfluency. Meanwhile there was little difference between those that did and did not have biological confirmation of their COVID-19 infection. This strongly suggests that self-reported cognitive deficits reported in our sample are associated with COVID-19 infection, rather than the experience of illness, or pandemic more generally.

### 4.3 Strengths, Limitations and Future Research

While the findings of this study are notable, there are a number of limitations in design and execution which warrant caution in interpreting the results.

First, this was an online study. Using online data-collection means that we are less able to maximize data quality by ensuring that participants were in a suitable environment or concentrating properly on the questionnaires. We were also not able to clinically assess participants, nor did we have access to medical records. This means that we were reliant on retrospective self-report for symptoms and diagnoses experienced sometimes months previously. In an attempt to reflect the feedback that we received from support groups during qualitative scoping, we used a slightly different symptom list when individuals were reporting on initial symptoms rather than ongoing symptoms, and the latter also had a greater range of possible values (reflecting both severity and regularity). This made it difficult to directly compare symptom profiles at the different time points, and future studies should consider using the same symptom list and reporting method for all time points, even if some symptoms are unlikely to appear at a given stage of illness. We also used a binary present/absent reporting approach for currently experienced symptoms, which was not able to reflect severity—this should also be addressed in future studies. To look at symptom profiles in terms of current symptoms, we used a *sum scores by factor* method (Hair 2009; Tabachnick et al. 2007) to calculate alignment of currently experienced symptoms with the symptom factors we established during the ongoing period. This was done for two reasons, partly because binary data does not lend itself to factor analysis, and partly because we believe that the co-occurrence of symptoms over a long time period (i.e., the “since then” ongoing period) is likely to be more reflective of underlying mechanism than the co-occurrence on a given day. As such, we believed it would be more informative to maintain the factors established during the longer period and use these as a reference for current illness. However, more detailed reporting of current symptoms would facilitate better interrogation of this data.

Much of the analysis in this study was necessarily exploratory, as too little was known at the time of study design to form many clear hypotheses. To handle this, multiple comparisons were conducted, for which the alpha adjustments entailed that only the very strongest effects survived at conventional statistical thresholds. This high type 2 error rate means that it is likely that more of the findings than just those that are reported here as statistically robust would be confirmed on replication. We have additionally reported the uncorrected results for this reason. A stated aim of this study was to generate hypotheses that could be tested in later, more targeted research, and we believe that for this purpose, many of the smaller effects that did not survive alpha correction may be worthy of further study.

We specifically targeted our recruitment to those self-identifying as suffering Long COVID, and we furthermore advertised the study as investigating memory and cognition in this group. As such, it is likely that our sample may have been biased towards those individuals with longer and more severe symptoms (who may be more likely to both seek a support group, and to be motivated to participate in studies in this area), and those experiencing cognitive symptoms (as these individuals may be more motivated to take part in a study explicitly investigating cognition). This study was not designed to assess prevalence, and as such it is not a major problem that Long COVID sufferers are over-represented relative to those that recovered. However, our reported rates of cognitive symptoms within the Long COVID cohort should be treated with caution, as they may represent an overestimation of true prevalence here. It is reassuring, however, that the figures for these symptoms within our cohort are comparable to those seen in other studies not explicitly investigating cognition (e.g., Davis et al. 2021; Ziauddeen et al. 2021).

Characterizing the sample, we found that those who had experienced COVID-19 infection were slightly older and more educated, and more likely to work in healthcare than those that had not experienced infection, who were more likely to be students. Those with severe ongoing symptoms were more likely to be older and more educated. We do not believe that these features reflect vulnerabilities towards COVID-19 or Long COVID, but rather the biases in our recruitment and target populations. Where possible, we controlled for age, sex, education and country of residence, which should mitigate some of these biases, however these sampling discrepancies should be kept in mind.

The original design of the study was to use the “No COVID” group as the basis for a before/after infection within-subjects comparison. As such, these participants were not in all ways treated as a matched comparison to the COVID group—in particular, they were not asked the same questions about experienced symptoms as the COVID group. This would have been highly useful in order to establish the degree to which symptoms (particularly those which might be expected to be exacerbated by lockdowns, such as depression, anxiety, fatigue) were more common in those that had previously experienced COVID-19 than those that had not. Moreover, those that reported having experienced COVID-19 but being fully recovered were not asked about current symptoms, making it difficult to compare to what extent these individuals differed from either the “No COVID” group or the “Ongoing symptoms group”. This is a design flaw in the study, which is addressed to some degree in follow-up sessions, where non-COVID participants are asked to report on symptoms.

However, we were able to compare symptoms in individuals who had experienced an illness that they did not think was COVID-19 to those that had had confirmed or unconfirmed COVID-19. This analysis found that a large number of symptoms were significantly more prevalent in those that had had suspected or confirmed COVID-19 infection compared to those who had experienced another (suspected non-COVID) illness during the same period.

### 4.4 Summary

The COVID and Cognition study is a cross-sectional/longitudinal study assessing symptoms, experiences and cognition in those that have experience COVID-19 infection. Here we present the first analysis in this cohort, characterizing the sample and investigating symptom profiles and cognitive symptoms in particular. We find that particular symptom-profiles—particularly neurological symptoms—during both the acute infection and ongoing illness were predictive of experience of cognitive dysfunction. The symptoms and experiences reported by our sample closely resemble those reported in previous work on Long COVID (e.g., Davis et al. 2021; Ziauddeen et al. 2021) which suggests that we can consider our, smaller, sample to be generally reflective of the larger Long COVID patient community. The participants in this study are being followed up over the course of the next 1–2 years, and it is hoped that future publications with this sample will provide valuable information as to the time-course of this illness.

The severity of the impact of “Long COVID” on everyday function and employment reported in our sample reflected previous studies (e.g., Davis et al. 2021) and is notable, particularly given the large proportion of healthcare and education staff in our sample. All of these issues should be of interest to policy makers, particularly when considering the extent to which large case numbers should be a concern in the context of reduced hospitalizations and deaths due to vaccination. While we do not yet know the impact of vaccination on Long COVID numbers, there are reasons to believe that high levels of infection among relatively young, otherwise healthy individuals may translate into considerable long-term workforce morbidity.

## Data Availability

All data produced in the present study are available upon reasonable request to the authors, and will be made available online soon.

## Acknowledgments

The COVID and Cognition study has benefitted from help and support from a large number of individuals, not least the participants who gave their time. We would especially like to thank members of the Long COVID support group, particularly Claire Hastie, Barbara Melville-Johannesson and Talya Varga for your time and insights. Thanks also to Emma Weisblatt, Mirjana Bozic and Keir Shiels who gave valuable suggestions, to Honor Thompson, Aashna Malik, Connor Doyle, Abel Ashby, Seraphina Zhang for research support and to Helen Spencer for statistical assistance. This study was not supported by any funding bodies but did benefit from research funds from the University of Cambridge department of Psychology.

