## Supplementary Materials for "COVCOG 1: Factors predicting Cognitive Symptoms in Long COVID. A First Publication from the COVID and Cognition Study"

**Supplementary Table 1.** Distribution of profession and employment status before the pandemic in No COVID and COVID groups. Note that participants could choose multiple options.

| | **No COVID (*n* = 185)** | **COVID (*n* = 181)** | **Chi-square tests** |
|---|---|---|---|
| Profession | | | |
| Not in paid work | 66 (35.7%) | 22 (12.2%) | $\chi^2(1) = 27.72$, $p < .001$, $V = .275$* |
| Healthcare setting | 16 (8.6%) | 40 (22.1%) | $\chi^2(1) = 12.77$, $p < .001$, $V = .187$* |
| Professional / Consultant / Academic | 26 (14.1%) | 29 (16%) | n.s. |
| Office Support / Administrative Work | 17 (9.2%) | 24 (13.3%) | n.s. |
| Education (Teaching / Childcare) | 7 (3.8%) | 19 (10.5%) | $\chi^2(1) = 6.25$, $p = .012$, $V = .131$ |
| Sales / Service | 13 (7%) | 8 (4.4%) | n.s. |
| Technician / Engineer / Mechanic | 2 (1.1%) | 7 (3.9%) | n.s. |
| Cleaner / Canteen Staff / Handyman | 7 (3.8%) | 1 (0.6%) | n.s. |

| | | | |
|---|---|---|---|
| Musician / Actor / Dancer and related performance work | 3 (1.6%) | 3 (1.7%) | n.s. |
| Construction / Agricultural / Forestry / Fishery worker | 5 (2.7%) | 1 (0.6%) | n.s. |
| Transport worker | 4 (2.2%) | 2 (1.1%) | n.s. |
| Craft / Art and related trade work | 3 (1.6%) | 1 (0.6%) | n.s. |
| Managerial | 1 (0.5%) | 1 (0.6%) | n.s. |
| Professional sports | - | 1 (0.6%) | n.s. |
| Other | 16 (14.1%) | 25 (13.8%) | n.s. |
| Employment status before pandemic | | | |
| Full time | 61 (33%) | 103 (56.9%) | $\chi^2(1) = 21.19$, $p < .001$, $V = .241$* |
| Part time | 39 (21.1%) | 39 (21.5%) | n.s. |
| Part time in multiple jobs | 6 (3.2%) | 6 (3.3%) | n.s. |
| Other schedule (e.g., ad hoc / gig economy) | 3 (1.6%) | 7 (3.9%) | n.s. |
| Not working (unemployed / career break / retired) | 42 (22.7%) | 16 (8.8%) | $\chi^2(1) = 13.18$, $p < .001$, $V = .190$* |
| Unpaid carer (e.g., children / elders) | 7 (3.8%) | 7 (3.9%) | n.s. |
| Student (e.g., University) | 50 (27%) | 26 (14.4%) | $\chi^2(1) = 8.91$, $p = .003$, $V = .156$* |

* denotes *p* values below Sidak-correct alpha (i.e. non-null)

**Supplementary Table 2.** Distribution of medical history in No COVID and COVID groups (top). Descriptive statistics and analyses for health behaviours factors (1 = *Never* – 6 = *Several times daily*; higher scores indicating higher frequency) in No COVID and COVID groups (bottom).

| | **No COVID** (*n* = 185) | **COVID** (*n* = 181) | |
|---|---|---|---|
| Medical history: Frequency (%) | | | **Chi-square tests** |
| Asthma | 35 (18.9%) | 38 (21%) | n.s. |
| Depression | 38 (20.5%) | 34 (18.8%) | n.s. |
| Other mental health disorder | 35 (18.9%) | 28 (15.5%) | n.s. |
| Obesity | 15 (8.1%) | 23 (12.7%) | n.s. |
| High blood pressure | 20 (10.8%) | 19 (10.5%) | n.s. |
| History of migraines | 7 (3.8%) | 19 (10.5%) | $\chi^2(1) = 6.25$, $p = .012$, $V = .131$ |
| Inflammatory / Autoimmune | 4 (2.2%) | 18 (9.9%) | $\chi^2(1) = 9.81$, $p = .002$, $V = .164$* |
| Chronic fatigue syndrome / Myalgic encephalomyelitis (ME) | 9 (4.9%) | 8 (4.4%) | n.s. |
| Psychiatric/neurodevelopmental disorder | 6 (3.2%) | 7 (3.9%) | n.s. |
| Cardiovascular disease / Angina | 4 (2.2%) | 6 (3.3%) | n.s. |
| Diabetes (Type 2) | 9 (4.9%) | 2 (1.1%) | n.s. |
| Diabetes (Type 1) | 4 (2.2%) | - | n.s. |
| Cancer | 1 (0.5%) | 2 (1.1%) | n.s. |
| A clotting disorder | 1 (0.5%) | 2 (1.1%) | n.s. |

| | | | |
|---|---|---|---|
| None of the above | 79 (42.7%) | 65 (35.9%) | n.s. |
| Health behaviours: *Mean (SD)* | | | **Mann-Whitney U tests** |
| Diet: Fruit and vegetables | 4.66 (1.16) | 5.04 (1.1) | COVID > No COVID: *U* = 13525, *p* = .001* |
| Diet: Sugary food | 3.69 (1.11) | 3.4 (1.07) | No COVID > COVID: *U* = 14168, *p* = .008* |
| Diet: Fatty food | 3.26 (1.01) | 3.06 (.86) | No COVID > COVID: *U* = 14825, *p* = .041 |
| Physical activity | 3.3 (1.24) | 3.7 (1.33) | COVID > No COVID: *U* = 13752, *p* = .002* |
| Alcohol | 2.67 (1.23) | 2.69 (1.02) | n.s. |
| Smoking | 1.63 (1.54) | 1.4 (1.19) | n.s. |

* denotes *p* values below Sidak-correct alpha

**Supplementary Table 3.** Distribution of demographics in COVID subgroups: Recovered (*R*), Ongoing (Mild/Moderate (*C+*)) and Ongoing (Severe (*C++*)).

| | **Recovered (*R*) (*n* = 42)** | **Ongoing (Mild/Moderate) (*C+*) (*n* = 53)** | **Ongoing (Severe) (*C++*) (*n* = 66)** | **Chi-square tests** |
|---|---|---|---|---|
| Weeks since infection | | | | |
| 3–10 | 15 (35.7%) | 7 (13.2%) | 4 (6.1%) | $\chi^2(6) = 19.64$, $p = .003$, |
| 11–20 | 1 (2.4%) | 4 (7.5%) | 5 (7.6%) | $V = .247$ |
| 21–30 | 12 (28.6%) | 15 (28.3%) | 17 (25.8%) | |
| 31+ | 14 (33.3%) | 27 (50.9%) | 40 (60.6%) | |
| Sex | | | | |
| Man | 13 (31%) | 16 (30.2%) | 10 (15.2%) | n.s. |
| Woman | 29 (69%) | 36 (67.9%) | 54 (81.8%) | |
| Other | - | 1 (1.9%) | 2 (3%) | |
| Age | | | | |
| 18-20 | 9 (21.4%) | 2 (3.8%) | - | $\chi^2(10) = 53.41$, $p < .001$, |
| 21-30 | 14 (33.3%) | 8 (15.1%) | 2 (3%) | $V = .407$ |
| 31-40 | 10 (23.8%) | 11 (20.8%) | 14 (21.2%) | |
| 41-50 | 3 (7.1%) | 14 (26.4%) | 16 (24.2%) | |
| 51-60 | 4 (9.5%) | 9 (17%) | 26 (39.4%) | |
| 61 or above | 2 (4.8%) | 9 (17%) | 8 (12.1%) | |
| Education | | | | |
| GCSE/equiv or below | 7 (16.7%) | 4 (7.5%) | 2 (3%) | |

| | | | | |
|---|---|---|---|---|
| A level/equiv | 7 (16.7%) | 4 (7.5%) | 2 (3%) | |
| Attended college (no degree) / Technical training / Assoc. degree | 7 (16.7%) | 13 (24.5%) | 8 (12.1%) | $\chi^2(10) = 20.03$, $p = .029$, |
| Bachelor's degree | 8 (19%) | 15 (28.3%) | 26 (39.4%) | $V = .249$ |
| Master's / Professional degree | 11 (26.2%) | 13 (24.5%) | 24 (36.4%) | |
| Doctorate degree | 2 (4.8%) | 4 (7.5%) | 4 (6.1%) | |
| Country | | | | |
| UK | 23 (54.8%) | 34 (64.2%) | 61 (92.4%) | $\chi^2(4) = 24.24$, $p < .001$, |
| North America | 10 (23.8%) | 12 (22.6%) | 5 (7.6%) | $V = .274$ |
| Other | 9 (21.4%) | 7 (13.2%) | - | |
| Ethnicity | | | | |
| Northern European | 29 (69%) | 48 (90.6%) | 62 (93.9%) | $\chi^2(2) = 14.68$, $p = .001$, $V = .302$ |
| Southern European / Latinx | 6 (14.3%) | 5 (9.4%) | 5 (7.5%) | n.s. |
| African / Afro-Caribbean | 4 (9.5%) | 1 (1.9%) | 2 (3%) | n.s. |
| Asian | 4 (9.6%) | 3 (5.7%) | - | n.s. |
| Other / Prefer not to say | 3 (7.2%) | 1 (1.9%) | 1 (1.5%) | n.s. |

**Supplementary Table 4.** Diagnoses during initial illness among those seeking medical attention ($N$ = 109).

| Diagnoses within initial illness period | Percentage of positive diagnosis (number) |
|---|---|
| **Hypoxia** | 14.7% (16) |
| **TIA (Transient ischemic attack)** | 0.9% (1) |
| **Clotting / Blood clots** | 5.5% (6) |
| **Encephalopathies** | 0.9% (1) |
| **Aneurism** | 1.8% (2) |
| **Inflammatory Syndrome** | 4.6% (5) |
| **Encephalitis** | 1.8% (2) |
| **Hematoma** | 0.9% (1) |
| **Kidney failure** | 0.9% (1) |
| **Encephalomyelitis** | - |
| **Respiratory arrest** | - |
| **Delirium** | 1.8% (2) |
| **Cardiac arrest** | 1.8% (2) |
| **Stroke** | 2.8% (3) |
| **Shock** | 1.8% (2) |

**Supplementary Table 5.** Descriptive statistics and analyses for initial symptom experience (recoded such that higher numbers indicate increased severity: 1 = *Not at all*, 3 = *Very severe*) in COVID subgroups: Recovered (*R*), Ongoing (Mild/Moderate (*C+*)) and Ongoing (Severe (*C++*)).

| | **Recovered (*R*) (*n* = 42)** | **Ongoing (Mild/Moderate) (*C+*) (*n* = 53)** | **Ongoing (Severe) (*C++*) (*n* = 66)** | **Kruskal-Wallis H tests** |
|---|---|---|---|---|
| Initial symptoms: *Mean (SD)* | | | | |
| Fatigue | 2.24 (.69) | 2.58 (.57) | 2.76 (.47) | $H(2) = 18.45, p < .001$* |
| Muscle/body pains | 1.81 (.67) | 2.17 (.58) | 2.44 (61) | $H(2) = 22.63, p < .001$* |
| Breathing issues | 1.62 (.73) | 2.04 (.73) | 2.44 (.7) | $H(2) = 28.04, p < .001$* |
| Headache | 2 (.73) | 2.04 (.68) | 2.35 (.71) | $H(2) = 8.51, p = .014$ |
| Brain fog | 1.62 (.76) | 2.17 (.7) | 2.26 (.79) | $H(2) = 17.67, p < .001$* |
| Chest pain/tightness | 1.55 (.63) | 2.13 (.65) | 2.26 (.73) | $H(2) = 25.22, p < .001$* |
| Limb weakness | 1.43 (.59) | 1.75 (.62) | 2.17 (.65) | $H(2) = 31.14, p < .001$* |
| Dizziness | 1.79 (.72) | 1.83 (.7) | 2.15 (.69) | $H(2) = 9.01, p = .011$ |
| Drowsiness | 1.88 (.74) | 2.19 (.71) | 2.14 (.72) | n.s. |
| Cough | 2.02 (.75) | 1.77 (.75) | 2.08 (.83) | n.s. |
| Loss of appetite | 1.74 (.70) | 1.7 (.75) | 2.03 (.78) | $H(2) = 6.51, p = .039$ |
| Fever | 1.71 (.64) | 1.7 (.77) | 1.98 (.79) | n.s. |
| Abdominal pain | 1.36 (.53) | 1.58 (.66) | 1.91 (.78) | $H(2) = 14.84, p = .001$* |
| Altered consciousness | 1.43 (.63) | 1.66 (.62) | 1.91 (.72) | $H(2) = 13.09, p = .001$* |
| Loss of smell/taste | 1.79 (.87) | 1.74 (.88) | 1.88 (.85) | n.s. |
| Sore throat | 1.67 (.69) | 1.75 (.65) | 1.85 (.69) | n.s. |

| | | | | |
|---|---|---|---|---|
| Nausea | 1.38 (.58) | 1.51 (.61) | 1.85 (.69) | $H(2) = 15.03$, $p = .001$* |
| Confusion | 1.33 (.61) | 1.58 (.69) | 1.83 (.74) | $H(2) = 13.94$, $p = .001$* |
| Eye-soreness | 1.48 (.67) | 1.6 (.63) | 1.8 (.73) | $H(2) = 6.25$, $p = .044$ |
| Diarrhea | 1.5 (.67) | 1.55 (.61) | 1.76 (.77) | n.s. |
| Anxiety | 1.62 (.7) | 1.72 (.63) | 1.68 (.71) | n.s. |
| Blurred vision | 1.19 (.46) | 1.32 (.55) | 1.62 (.67) | $H(2) = 15.45$, $p < .001$* |
| Disorientation | 1.17 (.38) | 1.47 (.58) | 1.48 (.66) | $H(2) = 8.78$, $p = .012$ |
| Acid reflux | 1.21 (.47) | 1.32 (.55) | 1.45 (.66) | n.s. |
| Visual disturbances | 1.19 (.4) | 1.19 (.44) | 1.36 (.65) | n.s. |
| Rash | 1.14 (.42) | 1.36 (.59) | 1.32 (.53) | n.s. |
| Speech difficulty | 1.14 (.35) | 1.26 (.45) | 1.32 (.53) | n.s. |
| Numbness | 1.05 (.22) | 1.23 (.47) | 1.3 (.53) | $H(2) = 8.51$, $p = .014$ |
| Vomiting | 1.17 (.44) | 1.08 (.33) | 1.23 (.46) | n.s. |
| Hallucinations | 1.07 (.26) | 1.08 (.27) | 1.23 (.55) | n.s. |
| Foot sores | 1.07 (.34) | 1.11 (.32) | 1.2 (.4) | n.s. |
| Itchy welts | 1.14 (.42) | 1.25 (.43) | 1.17 (.41) | n.s. |
| Delirium | 1.14 (.35) | 1.11 (.42) | 1.15 (.47) | n.s. |

** denotes p values below Sidak-correct alpha (i.e. non-null)*

**Supplementary Table 6.** Descriptive statistics and analyses for initial symptom experience (recoded such that higher numbers indicate increased severity: 1 = *Not at all*, 3 = *Very severe*) in Unknown, Unconfirmed COVID, and Confirmed COVID groups.

| | **Unknown** (*n* = 55) | **Unconfirmed** (*n* = 96) | **Confirmed** (*n* = 65) | **Kruskal-Wallis H tests** |
|---|---|---|---|---|
| Initial symptoms: *Mean (SD)* | | | | |
| Fatigue | 1.84 (.79) | 2.58 (.63) | 2.54 (.56) | $H(2) = 36.94, p < .001$* |
| Muscle/body pains | 1.58 (.63) | 2.17 (.71) | 2.22 (.6) | $H(2) = 30.09, p < .001$* |
| Loss of smell/taste | 1.22 (.57) | 1.63 (.77) | 2.08 (.92) | $H(2) = 30.92, p < .001$* |
| Drowsiness | 1.55 (.66) | 2.11 (.72) | 2.05 (.74) | $H(2) = 22, p < .001$* |
| Brain fog | 1.42 (.63) | 2.08 (.77) | 2.03 (.83) | $H(2) = 26.67, p < .001$* |
| Headache | 1.96 (.61) | 2.25 (.68) | 2.02 (.76) | $H(2) = 7.63, p = .022$ |
| Breathing issues | 1.51 (.66) | 2.22 (.76) | 1.91 (.81) | $H(2) = 27.87, p < .001$* |
| Dizziness | 1.53 (.6) | 2 (.7) | 1.88 (.74) | $H(2) = 15.78, p < .001$* |
| Chest pain/tightness | 1.4 (.56) | 2.16 (.73) | 1.85 (.71) | $H(2) = 36.05, p < .001$* |
| Fever | 1.49 (.64) | 1.8 (.73) | 1.85 (.8) | $H(2) = 7.96, p = .019$ |
| Loss of appetite | 1.45 (.66) | 1.86 (.73) | 1.82 (.81) | $H(2) = 11.72, p = .003$ |
| Limb weakness | 1.29 (.5) | 1.86 (.71) | 1.8 (.67) | $H(2) = 27.62, p < .001$* |
| Cough | 2.05 (.65) | 2.09 (.78) | 1.77 (.77) | $H(2) = 7.77, p = .021$ |
| Altered consciousness | 1.35 (.62) | 1.73 (.7) | 1.66 (.67) | $H(2) = 13.15, p = .001$* |
| Anxiety | 1.84 (.76) | 1.69 (.62) | 1.66 (.76) | n.s. |
| Eye-soreness | 1.38 (.56) | 1.69 (.65) | 1.6 (.75) | $H(2) = 7.89, p = .019$ |
| Sore throat | 2.02 (.62) | 1.9 (.67) | 1.58 (.64) | $H(2) = 14.61, p = .001$* |
| Diarrhea | 1.36 (.49) | 1.65 (.71) | 1.58 (.68) | n.s. |

| | | | | |
|---|---|---|---|---|
| Abdominal pain | 1.4 (.6) | 1.72 (.72) | 1.57 (.71) | *H*(2) = 7.54, *p* = .023 |
| Nausea | 1.38 (.53) | 1.67 (.66) | 1.54 (.66) | *H*(2) = 6.78, *p* = .034 |
| Confusion | 1.25 (.52) | 1.69 (.74) | 1.52 (.67) | *H*(2) = 14.15, *p* = .001* |
| Acid reflux | 1.22 (.46) | 1.29 (.54) | 1.43 (.64) | n.s. |
| Disorientation | 1.18 (.43) | 1.45 (.61) | 1.32 (.53) | *H*(2) = 8.37, *p* = .015 |
| Blurred vision | 1.2 (.52) | 1.5 (.62) | 1.28 (.57) | *H*(2) = 15.03, *p* = .001* |
| Rash | 1.18 (.51) | 1.3 (.53) | 1.26 (.54) | n.s. |
| Visual disturbances | 1.13 (.34) | 1.29 (.54) | 1.22 (.52) | n.s. |
| Numbness | 1.07 (.26) | 1.23 (.47) | 1.18 (.43) | n.s. |
| Speech difficulty | 1.04 (.19) | 1.3 (.51) | 1.18 (.39) | *H*(2) = 13.76, *p* = .001* |
| Itchy welts | 1.18 (.55) | 1.21 (.43) | 1.15 (.4) | n.s. |
| Foot sores | 1.04 (.27) | 1.15 (.38) | 1.12 (.33) | n.s. |
| Vomiting | 1.15 (.41) | 1.19 (.44) | 1.12 (.38) | n.s. |

** denotes p values below Sidak-correct alpha (i.e. non-null)*

**Supplementary Table 7.** Descriptive statistics and analyses for ongoing symptom experience (recoded such that higher numbers indicate increased severity: 1 = *Not at all*, 5 = *Very severe and often*) in Unknown, Unconfirmed COVID, and Confirmed COVID groups.

| | **Unknown (n = 31)** | **Unconfirmed (n = 88)** | **Confirmed (n = 58)** | **Kruskal-Wallis H tests** |
|---|---|---|---|---|
| Ongoing symptoms: *Mean (SD)* | | | | |
| Fatigue | 2.55 (1.5) | 3.77 (1.35) | 3.83 (1.39) | $H(2) = 16.86$, $p < .001$* |
| Difficulty concentrating | 2.03 (1.05) | 3.64 (1.22) | 3.52 (1.3) | $H(2) = 32.58$, $p < .001$* |
| Brain fog | 1.97 (1.33) | 3.43 (1.45) | 3.24 (1.49) | $H(2) = 20.73$, $p < .001$* |
| Forgetfulness | 2.35 (1.31) | 3.27 (1.32) | 3.19 (1.34) | $H(2) = 10.92$, $p = .004$ |
| Tip of the Tongue (ToT) | 1.65 (0.92) | 3 (1.41) | 3.16 (1.41) | $H(2) = 25.71$, $p < .001$* |
| Difficulty sleeping | 2.35 (1.33) | 3.15 (1.41) | 2.86 (1.16) | $H(2) = 8.09$, $p = .017$ |
| Muscle/body pains | 1.71 (0.78) | 2.83 (1.39) | 2.83 (1.48) | $H(2) = 16.32$, $p < .001$* |
| Breathing issues | 2 (1.29) | 2.98 (1.47) | 2.83 (1.4) | $H(2) = 11.07$, $p = .004$ |
| Fast/irregular pulse | 1.39 (0.88) | 2.85 (1.41) | 2.81 (1.5) | $H(2) = 29.07$, $p < .001$* |
| Semantic disfluency | 1.35 (0.88) | 2.64 (1.42) | 2.78 (1.49) | $H(2) = 26.08$, $p < .001$* |
| Night waking | 2.23 (1.33) | 3.24 (1.38) | 2.74 (1.33) | $H(2) = 13.18$, $p = .001$* |
| Chest pain/tightness | 1.84 (1.21) | 2.89 (1.44) | 2.69 (1.26) | $H(2) = 14.61$, $p = .001$* |
| Vivid dreams | 2.03 (1.3) | 2.78 (1.15) | 2.62 (1.32) | $H(2) = 9.12$, $p = .01$ |
| Dizziness | 1.9 (1.17) | 2.67 (1.32) | 2.57 (1.42) | $H(2) = 8.33$, $p = .016$ |
| Drowsiness | 2.06 (1.39) | 2.35 (1.25) | 2.53 (1.33) | n.s. |
| Headache | 2.32 (1.05) | 2.78 (1.33) | 2.53 (1.31) | n.s. |
| Limb weakness | 1.42 (0.67) | 2.38 (1.34) | 2.47 (1.47) | $H(2) = 14.53$, $p < .001$* |
| Depression | 3.03 (1.33) | 2.51 (1.3) | 2.43 (1.24) | n.s. |

| | | | | |
|---|---|---|---|---|
| Excess thirst | 1.77 (1.12) | 2.43 (1.35) | 2.41 (1.36) | $H(2) = 6.69$, $p = .035$ |
| Loss of smell/taste | 1.16 (0.52) | 1.84 (1.21) | 2.38 (1.62) | $H(2) = 16.25$, $p < .001$* |
| Head pressure | 1.68 (0.95) | 2.4 (1.38) | 2.31 (1.35) | $H(2) = 6.53$, $p = .038$ |
| Anxiety | 2.65 (1.52) | 2.45 (1.33) | 2.24 (1.26) | n.s. |
| Confusion | 1.39 (0.67) | 2.18 (1.3) | 2.22 (1.36) | $H(2) = 9.89$, $p = .007$ |
| Weight gain | 1.81 (1.25) | 2.1 (1.33) | 2.16 (1.35) | n.s. |
| Abdominal pain | 1.71 (1.07) | 2.22 (1.16) | 2 (1.14) | $H(2) = 6.44$, $p = .04$ |
| Pins & needles | 1.68 (1.01) | 2.19 (1.26) | 1.98 (1.16) | n.s. |
| Sore throat | 1.94 (1.03) | 2.33 (1.22) | 1.93 (1.23) | $H(2) = 6.38$, $p = .041$ |
| Loss of appetite | 1.48 (0.57) | 1.83 (1.08) | 1.9 (1.28) | n.s. |
| Altered consciousness | 1.65 (1.17) | 1.85 (1.17) | 1.9 (1.2) | n.s. |
| Diarrhea | 1.52 (0.81) | 1.91 (1.02) | 1.9 (1.14) | n.s. |
| Acid reflux | 1.39 (0.8) | 1.68 (1.09) | 1.88 (1.26) | n.s. |
| Eye-soreness | 1.65 (1.08) | 2.26 (1.19) | 1.88 (1.31) | $H(2) = 11.27$, $p = .004$ |
| Blurred vision | 1.39 (0.76) | 2.03 (1.24) | 1.84 (1.27) | $H(2) = 8.14$, $p = .017$ |
| Nausea | 1.48 (0.85) | 1.89 (0.95) | 1.79 (1.21) | $H(2) = 6.32$, $p = .042$ |
| Cough | 2.16 (1.39) | 2.31 (1.16) | 1.79 (0.95) | $H(2) = 7.12$, $p = .028$ |
| Hot flushes | 1.26 (0.58) | 1.84 (1.12) | 1.74 (1.21) | $H(2) = 7.88$, $p = .019$ |
| Weight loss | 1.48 (0.89) | 1.52 (0.99) | 1.62 (1.09) | n.s. |
| Disorientation | 1.29 (0.82) | 1.57 (0.84) | 1.48 (0.98) | n.s. |
| Visual disturbances | 1.23 (0.62) | 1.52 (0.98) | 1.47 (0.98) | n.s. |
| Speech difficulty | 1.19 (0.65) | 1.66 (0.98) | 1.43 (0.9) | $H(2) = 9.45$, $p = .009$ |
| Numbness | 1.23 (0.62) | 1.6 (1.06) | 1.43 (0.94) | n.s. |
| Rash | 1.39 (0.96) | 1.55 (1.04) | 1.4 (0.79) | n.s. |
| Fever | 1.13 (0.43) | 1.42 (0.85) | 1.33 (0.85) | n.s. |

| | | | | |
|---|---|---|---|---|
| Incontinence | 1.16 (0.58) | 1.27 (0.6) | 1.28 (0.7) | n.s. |
| Itchy welts | 1.32 (0.91) | 1.24 (0.66) | 1.22 (0.53) | n.s. |
| Foot sores | 1.06 (0.25) | 1.22 (0.65) | 1.17 (0.57) | n.s. |
| Vomiting | 1.13 (0.43) | 1.19 (0.48) | 1.17 (0.53) | n.s. |

** denotes p values below Sidak-correct alpha (i.e. non-null)*

**Supplementary Table 8.** Factors and loadings from the “since then” cognitive symptoms PCA.

| | **Cognitive Symptoms** |
|---|---|
| Brain Fog | 0.889 |
| Forgetfulness | 0.881 |
| Tip of the Tongue (ToT) | 0.879 |
| Semantic Disfluency | 0.874 |
| Difficulty Concentrating | 0.859 |

**Supplementary Table 9.** Distribution of experience with long COVID in COVID subgroups: Recovered (*R*), Ongoing (Mild/Moderate (*C+*)) and Ongoing (Severe (*C++*)).

| | **Now Recovered (*R*) (*n* = 27**)** | **Ongoing (Mild/Moderate) (*C+*) (*n* = 53)** | **Ongoing (Severe) (*C++*) (*n* = 66)** | **Chi-square tests** |
|---|---|---|---|---|
| Identify as suffering from "Long COVID" | | | | |
| Yes | 3 (11.1%) | 43 (81.1%) | 62 (93.9%) | $\chi^2(4) = 85.75$, $p < .001$, |
| No | 16 (59.3%) | 2 (3.8%) | - | $V = .542$ |
| Other | 8 (29.6%) | 8 (15.1%) | 4 (6.1%) | |
| Change of symptoms after initial illness | | | | |
| No ongoing symptoms after initial recovery | 5 (18.5%) | - | - | |
| Different symptoms at different times | 8 (29.6%) | 28 (52.8%) | 39 (59.1%) | $\chi^2(6) = 37.52$, $p < .001$, |
| Improvement in symptoms over time | 5 (18.5%) | 18 (34%) | 9 (13.6%) | $V = .367$ |
| Symptoms have been very consistent | 3 (11.1%) | 7 (13.2%) | 17 (25.8%) | |
| I don't know / N/A | 6 (22.2%) | - | 1 (1.5%) | |
| Cycle of symptoms after initial illness | | | | |
| Cycle every few days | 3 (11.1%) | 11 (20.8%) | 14 (21.2%) | |
| Cycle every few weeks | 3 (11.1%) | 13 (24.5%) | 19 (28.8%) | |
| Cycle monthly | 2 (7.4%) | 7 (13.2%) | 9 (13.6%) | n.s. |

| | | | | |
|---|---|---|---|---|
| No cycling | 12 (44.4%) | 19 (35.8%) | 23 (34.8%) | |
| N/A | 7 (25.9%) | 3 (5.7%) | 1 (1.5%) | |
| Impacts of long COVID | | | | |
| Long period unable to work (due to illness) | 2 (7.4%) | 15 (28.3%) | 50 (75.8%) | $\chi^2(2) = 46.42$, $p < .001$, $V = .564$* |
| Difficulty coping day-to-day activities | 6 (22.2%) | 28 (52.8%) | 48 (72.7%) | $\chi^2(2) = 20.23$, $p < .001$, $V = .372$* |
| Difficulty getting medical professionals to take symptoms seriously | 1 (3.7%) | 21 (39.6%) | 38 (57.6%) | $\chi^2(2) = 23.05$, $p < .001$, $V = .397$* |
| Lost job due to illness | 1 (3.7%) | 9 (17%) | 32 (48.5%) | $\chi^2(2) = 24.39$, $p < .001$, $V = .409$* |
| Feeling that you have experienced a trauma | 4 (14.8%) | 21 (39.6%) | 31 (47%) | $\chi^2(2) = 8.44$, $p = .015$, $V = .240$ |
| Financial difficulty (as a result of illness) | 1 (3.7%) | 7 (13.2%) | 14 (21.2%) | n.s. |
| None of the above | 18 (66.7%) | 9 (17%) | 1 (1.5%) | $\chi^2(2) = 52.73$, $p < .001$, $V = .601$* |

* denotes p values below Sidak-correct alpha at .007 for the impacts of long COVID

**Excluding a small portion of participants who reported asymptomatic or feeling completely better very quickly from the Recovered subgroup (*n* = 15)

**Supplementary Table 10.** Descriptive statistics and analyses for ongoing symptom experience (recoded such that higher numbers indicate increased severity: 1 = *Not at all*, 5 = *Very severe and often*) in COVID subgroups: Recovered (*R*), Ongoing (Mild/Moderate (*C+*)) and Ongoing (Severe (*C++*)).

| | **Now Recovered (*R*) (*n* = 27**)** | **Ongoing (Mild/Moderate) (*C+*) (*n* = 53)** | **Ongoing (Severe) (*C++*) (*n* = 66)** | **Kruskal-Wallis H tests** |
|---|---|---|---|---|
| Ongoing symptoms: *Mean (SD)* | | | | |
| Fatigue | 2.33 (1.24) | 3.57 (1.12) | 4.58 (0.98) | *H*(2) = 57.34, *p* < .001* |
| Difficulty concentrating | 2.33 (1.36) | 3.26 (1) | 4.36 (0.78) | *H*(2) = 52.98, *p* < .001* |
| Brain fog | 1.85 (1.26) | 3.17 (1.21) | 4.12 (1.18) | *H*(2) = 46.08, *p* < .001* |
| Forgetfulness | 2.07 (1.24) | 2.98 (1.08) | 3.92 (1.13) | *H*(2) = 39.24, *p* < .001* |
| Tip of the Tongue (ToT) | 1.78 (1.16) | 2.81 (1.18) | 3.79 (1.23) | *H*(2) = 41.03, *p* < .001* |
| Breathing issues | 1.63 (1.15) | 2.57 (1.07) | 3.73 (1.32) | *H*(2) = 46.08, *p* < .001* |
| Fast/irregular pulse | 1.59 (0.8) | 2.45 (1.25) | 3.65 (1.31) | *H*(2) = 45.11, *p* < .001* |
| Night waking | 2 (1.27) | 2.87 (1.35) | 3.61 (1.16) | *H*(2) = 27.25, *p* < .001* |
| Muscle/body pains | 1.67 (0.83) | 2.51 (1.15) | 3.56 (1.41) | *H*(2) = 37.66, *p* < .001* |
| Chest pain/tightness | 1.70 (1.03) | 2.43 (1.14) | 3.56 (1.24) | *H*(2) = 42.21, *p* < .001* |
| Semantic disfluency | 1.59 (0.84) | 2.19 (1.24) | 3.55 (1.3) | *H*(2) = 45.81, *p* < .001* |
| Difficulty sleeping | 2.67 (1.44) | 2.81 (1.21) | 3.36 (1.3) | *H*(2) = 7.47, *p* = .024 |
| Headache | 1.96 (1.19) | 2.23 (1.03) | 3.35 (1.3) | *H*(2) = 30.33, *p* < .001* |
| Dizziness | 1.81 (1.18) | 2.19 (1.04) | 3.32 (1.33) | *H*(2) = 31.94, *p* < .001* |
| Limb weakness | 1.22 (0.42) | 2.13 (1.09) | 3.12 (1.45) | *H*(2) = 40.54, *p* < .001* |
| Head pressure | 1.67 (0.92) | 1.94 (0.97) | 2.98 (1.52) | *H*(2) = 22.01, *p* < .001* |

| | | | | |
|---|---|---|---|---|
| Vivid dreams | 2.07 (1.17) | 2.79 (1.1) | 2.92 (1.26) | $H(2) = 9.94$, $p = .007$ |
| Excess thirst | 1.74 (1.16) | 2.23 (1.2) | 2.86 (1.40) | $H(2) = 15.74$, $p < .001$* |
| Sore throat | 1.33 (0.56) | 1.91 (0.9) | 2.73 (1.4) | $H(2) = 26.45$, $p < .001$* |
| Drowsiness | 1.78 (1.01) | 2.4 (1.28) | 2.71 (1.3) | $H(2) = 11.11$, $p = .004$ |
| Confusion | 1.33 (0.83) | 2.02 (1.01) | 2.7 (1.49) | $H(2) = 21.82$, $p < .001$* |
| Pins & needles | 1.41 (0.64) | 1.75 (0.88) | 2.68 (1.38) | $H(2) = 24.69$, $p < .001$* |
| Abdominal pain | 1.44 (0.7) | 1.81 (0.74) | 2.67 (1.33) | $H(2) = 25.12$, $p < .001$* |
| Eye-soreness | 1.48 (0.8) | 1.94 (1.08) | 2.5 (1.4) | $H(2) = 13.46$, $p = .001$* |
| Blurred vision | 1.19 (0.56) | 1.68 (0.98) | 2.5 (1.41) | $H(2) = 28.35$, $p < .001$* |
| Cough | 1.59 (0.84) | 1.92 (1.04) | 2.46 (1.16) | $H(2) = 14.55$, $p = .001$* |
| Anxiety | 2.19 (1.3) | 2.38 (1.35) | 2.44 (1.28) | n.s. |
| Depression | 2.48 (1.37) | 2.55 (1.2) | 2.42 (1.31) | n.s. |
| Weight gain | 1.56 (0.75) | 2.06 (1.17) | 2.41 (1.56) | n.s. |
| Loss of smell/taste | 1.63 (1.25) | 1.89 (1.2) | 2.36 (1.57) | $H(2) = 6.27$, $p = .044$ |
| Nausea | 1.22 (0.51) | 1.62 (0.74) | 2.29 (1.25) | $H(2) = 22.88$, $p < .001$* |
| Loss of appetite | 1.59 (0.97) | 1.51 (0.89) | 2.23 (1.31) | $H(2) = 14.11$, $p = .001$* |
| Diarrhoea | 1.44 (0.58) | 1.77 (0.89) | 2.2 (1.26) | $H(2) = 8.28$, $p = .016$ |
| Altered consciousness | 1.22 (0.51) | 1.81 (1.13) | 2.18 (1.3) | $H(2) = 15.09$, $p = .001$* |
| Hot flushes | 1.33 (0.78) | 1.68 (0.96) | 2.09 (1.34) | $H(2) = 8.66$, $p = .013$ |
| Acid reflux | 1.33 (0.62) | 1.68 (1.12) | 2 (1.3) | n.s. |
| Speech difficulty | 1.04 (0.19) | 1.38 (0.63) | 1.94 (1.19) | $H(2) = 18.85$, $p < .001$* |
| Numbness | 1.11 (0.32) | 1.36 (0.79) | 1.85 (1.24) | $H(2) = 11.93$, $p = .003$ |
| Visual disturbances | 1.15 (0.46) | 1.31 (0.7) | 1.8 (1.22) | $H(2) = 10.6$, $p = .005$ |
| Disorientation | 1.07 (0.27) | 1.47 (0.75) | 1.77 (1.08) | $H(2) = 11.79$, $p = .003$ |
| Rash | 1.07 (0.27) | 1.42 (0.93) | 1.71 (1.08) | $H(2) = 11.39$, $p = .003$ |

| | | | | |
|---|---|---|---|---|
| Fever | 1.15 (0.46) | 1.15 (0.41) | 1.67 (1.11) | *H*(2) = 11.63, *p* = .003 |
| Weight loss | 1.52 (1.05) | 1.51 (0.95) | 1.62 (1.09) | n.s. |
| Incontinence | 1.22 (0.58) | 1.25 (0.55) | 1.32 (0.73) | n.s. |
| Itchy welts on skin | 1.07 (0.27) | 1.25 (0.62) | 1.29 (0.7) | n.s. |
| Foot sores | 1.07 (0.39) | 1.15 (0.5) | 1.29 (0.76) | n.s. |
| Vomiting | 1.11 (0.42) | 1.09 (0.35) | 1.26 (0.62) | n.s. |

** denotes p values below Sidak-correct alpha (i.e. non-null)*

**Excluding a small portion of participants who reported asymptomatic or feeling completely better very quickly from the Recovered subgroup (*n* = 15)

Analysis on Current symptoms

We examined symptoms reported as present or absent over the past 1–2 days ($n$ = 124, see Supplementary Table 11). Twelve symptoms appeared in less than 10% participants so were not included in the analyses. Pearson's chi-square tests with Sidak correction adjusting the alpha level to .0013 showed that the COVID subgroups significantly differed in their current experience of semantic disfluency only, with the symptom appearing more commonly in the *C++* (Severe) subgroup than the other subgroups.

**Supplementary Table 11.** Distribution of current symptoms in COVID subgroups: Recovered (*R*), Ongoing (Mild/Moderate (*C+*)) and Ongoing (Severe (*C++*)).

| | **Recovered (*R*) (*n* = 8**)** | **Ongoing (Mild/Moderate) (*C+*) (*n* = 50**)** | **Ongoing (Severe) (*C++*) (*n* = 66)** | **Chi-square tests** |
|---|---|---|---|---|
| Current symptoms: Frequency (%) | | | | |
| Fatigue | 5 (62.5%) | 37 (74%) | 57 (86.4%) | n.s. |
| Difficulty concentrating | 5 (62.5%) | 36 (72%) | 56 (84.8%) | n.s. |
| Forgetfulness | 3 (37.5%) | 30 (60%) | 52 (78.8%) | $\chi^2(2) = 8.48$, $p = .014$, $V = .262$ |
| Brain fog | 2 (25%) | 34 (68%) | 50 (75.8%) | $\chi^2(2) = 8.72$, $p = .013$, $V = .265$ |
| Breathing issues | 2 (25%) | 24 (48%) | 49 (74.2%) | $\chi^2(2) = 12.7$, $p = .002$, $V = .32$ |
| Tip of the Tongue (ToT) | 1 (12.5%) | 28 (56%) | 46 (69.7%) | $\chi^2(2) = 10.47$, $p = .005$, $V = .291$ |

| | | | | |
|---|---|---|---|---|
| Night waking | 5 (62.5%) | 28 (56%) | 43 (65.2%) | n.s. |
| Chest pain/tightness | 1 (12.5%) | 24 (48%) | 43 (65.2%) | $\chi^2(2) = 9.57$, $p = .008$, $V = .278$ |
| Muscle/body pains | 2 (25%) | 22 (44%) | 43 (65.2%) | $\chi^2(2) = 8.03$, $p = .018$, $V = .254$ |
| Headache | 2 (25%) | 20 (40%) | 40 (60.6%) | $\chi^2(2) = 6.97$, $p = .031$, $V = .237$ |
| Semantic disfluency | 1 (12.5%) | 13 (26%) | 40 (60.6%) | $\chi^2(2) = 17.21$, $p < .001$, $V = .373$* |
| Dizziness | 4 (50%) | 14 (28%) | 38 (57.6%) | $\chi^2(2) = 10.13$, $p = .006$, $V = .286$ |
| Fast/irregular pulse | 2 (25%) | 17 (34%) | 35 (53%) | n.s. |
| Difficulty sleeping | 5 (62.5%) | 19 (38%) | 32 (48.5%) | n.s. |
| Head pressure | 1 (12.5%) | 15 (30%) | 32 (48.5%) | $\chi^2(2) = 6.57$, $p = .037$, $V = .23$ |
| Sore throat | - | 14 (28%) | 28 (42.4%) | $\chi^2(2) = 7.02$, $p = .03$, $V = .238$ |
| Eye-soreness | 2 (25%) | 15 (30%) | 27 (40.9%) | n.s. |
| Limb weakness | 1 (12.5%) | 16 (32%) | 27 (40.9%) | n.s. |
| Anxiety | 3 (37.5%) | 19 (38%) | 25 (37.9%) | n.s. |
| Blurred vision | 1 (12.5%) | 7 (14%) | 25 (37.9%) | $\chi^2(2) = 9.18$, $p = .01$, $V = .272$ |
| Vivid dreams | 2 (25%) | 18 (36%) | 24 (36.4%) | n.s. |
| Depression | 4 (50%) | 22 (44%) | 23 (34.8%) | n.s. |
| Weight gain | 2 (25%) | 13 (26%) | 22 (33.3%) | n.s. |

| | | | | |
|---|---|---|---|---|
| Pins & needles | 2 (25%) | 9 (18%) | 22 (33.3%) | n.s. |
| Cough | 2 (25%) | 15 (30%) | 21 (31.8%) | n.s. |
| Abdominal pain | - | 9 (18%) | 21 (31.8%) | n.s. |
| Loss of smell/taste | 1 (12.5%) | 14 (28%) | 19 (28.8%) | n.s. |
| Confusion | 2 (25%) | 9 (18%) | 19 (28.8%) | n.s. |
| Excess thirst | 6 (75%) | 17 (34%) | 18 (27.3%) | $\chi^2(2) = 7.38$, $p = .025$, $V = .244$ |
| Hot flushes | 2 (25%) | 6 (12%) | 18 (27.3%) | n.s. |
| Drowsiness | 2 (25%) | 14 (28%) | 18 (27.3%) | n.s. |
| Nausea | - | 8 (16%) | 16 (24.2%) | n.s. |
| Loss of appetite | - | 7 (14%) | 13 (19.7%) | n.s. |
| Speech difficulty | - | 1 (2%) | 13 (19.7%) | $\chi^2(2) = 9.98$, $p = .007$, $V = .284$ |
| Diarrhoea | 1 (12.5%) | 11 (22%) | 13 (19.7%) | n.s. |
| Altered consciousness | - | 7 (14%) | 12 (18.2%) | n.s. |
| Acid reflux | - | 8 (16%) | 12 (18.2%) | n.s. |
| Numbness | - | 3 (6%) | 11 (16.7%) | n.s. |
| Weight loss | 1 (12.5%) | 7 (14%) | 9 (13.6%) | n.s. |
| Visual disturbances | - | 4 (8%) | 9 (13.6%) | n.s. |

** denotes p values below Sidak-correct alpha (i.e. non-null)*

**Further excluding a small portion of participants who reported asymptomatic or feeling completely better very quickly from the Recovered subgroup (*n* = 15) and an additional group of participants who reported not experiencing current symptoms (*n* = 22)

**Supplementary Table 12.** Factor loadings for current symptoms (binary present/absent data).

| Symptom | Component | | | |
|---|---|---|---|---|
| | **1** | **2** | **3** | **4** |
| Breathing issues | **0.647** | | | |
| Chest pain/tightness | **0.593** | | | |
| Fatigue | **0.557** | 0.302 | | |
| Weight gain | **0.534** | | | |
| Cough | **0.533** | | | |
| Muscle/body pains | **0.513** | 0.3 | | |
| Sore throat | **0.508** | | | |
| Night waking | **0.488** | | 0.337 | |
| Vivid dreams | **0.386** | | | |
| Excess thirst | **0.368** | | | |
| Fever | **0.352** | | | |
| Loss of smell/taste | **0.294** | | | |
| Rash | **0.254** | | | |
| Vomiting | **0.263** | | | |
| Hot flushes | | **0.585** | | |
| Abdominal pain | | **0.584** | | |
| Pins & needles | | **0.513** | | 0.317 |
| Eye-soreness | | **0.489** | | |
| Fast/irregular pulse | 0.412 | **0.473** | | |
| Headache | | **0.472** | 0.397 | |
| Face/lips swelling | | **0.455** | | |
| Dizziness | | **0.447** | 0.32 | |
| Blurred vision | | **0.446** | | 0.343 |
| Nausea | | **0.44** | | |
| Foot sores | | **0.425** | | |
| Head pressure | | **0.413** | 0.374 | |
| Itchy welts | | **0.296** | | |
| Diarrhea | | **0.291** | | |
| Weight loss | | | **0.684** | |
| Loss of appetite | | | **0.637** | |
| Drowsiness | | | **0.557** | |

| | | | |
|---|---|---|---|
| Speech difficulty | | **0.554** | 0.316 |
| Confusion | | **0.553** | |
| Depression | 0.423 | **0.46** | |
| Difficulty sleeping | | **0.418** | |
| Anxiety | 0.375 | **0.389** | |
| Delirium | | | **0.741** |
| Disorientation | | | **0.737** |
| Hallucinations | | | **0.696** |
| Visual disturbances | | | **0.51** |
| Altered consciousness | | | **0.446** |
